# Plasma Markers of Disrupted Gut Permeability in Severe COVID-19 Patients

**DOI:** 10.1101/2020.11.13.20231209

**Authors:** Leila B. Giron, Harsh Dweep, Xiangfan Yin, Han Wang, Mohammad Damra, Aaron R. Goldman, Nicole Gorman, Clovis S. Palmer, Hsin-Yao Tang, Maliha W. Shaikh, Christopher B. Forsyth, Robert A. Balk, Netanel F Zilberstein, Qin Liu, Andrew Kossenkov, Ali Keshavarzian, Alan Landay, Mohamed Abdel-Mohsen

## Abstract

A disruption of the crosstalk between the gut and the lung has been implicated as a driver of severity during respiratory-related diseases. Lung injury causes systemic inflammation, which disrupts gut barrier integrity, increasing the permeability to gut microbes and their products. This exacerbates inflammation, resulting in positive feedback. We aimed to test whether severe Coronavirus disease 2019 (COVID-19) is associated with markers of disrupted gut permeability. We applied a multi-omic systems biology approach to analyze plasma samples from COVID-19 patients with varying disease severity and SARS-CoV-2 negative controls. We investigated the potential links between plasma markers of gut barrier integrity, microbial translocation, systemic inflammation, metabolome, lipidome, and glycome, and COVID-19 severity. We found that severe COVID-19 is associated with high levels of markers of tight junction permeability and translocation of bacterial and fungal products into the blood. These markers of disrupted intestinal barrier integrity and microbial translocation correlate strongly with higher levels of markers of systemic inflammation and immune activation, lower levels of markers of intestinal function, disrupted plasma metabolome and glycome, and higher mortality rate. Our study highlights an underappreciated factor with significant clinical implications, disruption in gut functions, as a potential force that may contribute to COVID-19 severity.

## INTRODUCTION

Coronavirus Disease 2019 (COVID-19), the disease caused by severe acute respiratory syndrome coronavirus 2 (SARS-CoV-2) infection, can manifest with diverse clinical presentations. While the majority of infected individuals exhibit asymptomatic or mild respiratory tract infection, a significant population face severe manifestations such as acute respiratory distress syndrome (ARDS), multi-organ failure, and death (1). A state of hyper-inflammation and hyperactivated immune responses, characterized by an ensuing cytokine storm and increased complement activation, has been associated with COVID-19 severity (1, 2). However, the pathophysiological mechanisms that contribute to these phenomena remain mostly unknown. Understanding these mechanisms is a crucial step in designing rational clinical and therapeutic strategies.

A disruption of the crosstalk between the gut and the lung has been implicated as a driver of severity during respiratory-related diseases, including ARDS (3, 4). Systemic inflammation caused by a lung infection or injury can lead to a disruption of the gut barrier integrity and increase the permeability to gut microbes and microbial products. This microbial translocation can exacerbate systemic inflammation and lung injury – resulting in positive feedback (3, 4). In addition, SARS-CoV-2 can directly infect gut cells (5), and viral infections of the gut cause changes in gut structure and breakdown of the epithelial barrier (6).

Even as microbial translocation impacts systemic inflammation directly, it may also impact it indirectly by modulating circulating levels of gut- and gut microbiota-associated products such as metabolites and lipids. Plasma metabolites and lipids can reflect the functional status of the gut and the metabolic activity of its microbiota (7). They also are biologically active molecules in their own right, regulating several immunological functions, including inflammatory responses (8). A third class of microbial products that can translocate from the gut is glycan-degrading enzymes. Glycans on circulating glycoproteins and antibodies (IgGs and IgAs) are essential for regulating several immunological responses, including complement activation (9). The glycan-degrading enzymes are released by several members of the gut microbiome and their translocation can alter the circulating glycome, leading to higher inflammation and complement activation (10). Indeed, altered glycosylation of plasma glycoproteins (including immunoglobulin G, IgG) has been associated with the onset and progression of inflammatory bowel disease (IBD) (11). Furthermore, modulation of the gut microbiota via fecal microbiota transplantation affects IgG and serum glycosylation (12).

Here, we aimed to examine whether severe COVID-19 is associated with plasma markers of disrupted gut functions. Towards this aim, we applied a multi-omic systems biology approach to analyze plasma samples from COVID-19 patients with varying disease severity and SARS-CoV-2 negative controls.

## MATERIALS AND METHODS

### Study main cohort

Main analyses were performed using plasma samples from 60 individuals tested positive for SARS-CoV-2 and 20 SARS-CoV-2 negative controls collected at Rush University Medical Center (RUMC). The 60 SARS-CoV-2 positives were selected to represent three disease states: 20 with mild symptoms (outpatients); 20 with moderate symptoms (inpatients hospitalized on regular wards); and 20 with severe symptoms (inpatients hospitalized in an intensive care unit (ICU)) (**Figure 1a**). Individuals were selected to have a median age between 52.5 to 58.5 years. There was no significant difference in age between groups (**Supplementary Figure 1**). The study cohort was also chosen to have a 35 to 60% representation of female gender per disease status group (**Supplementary Table 1**). Eight participants of the cohort (two from the moderate group and six from the severe group) died from COVID-19 (**Supplementary Table 1**).

**Figure 1.**
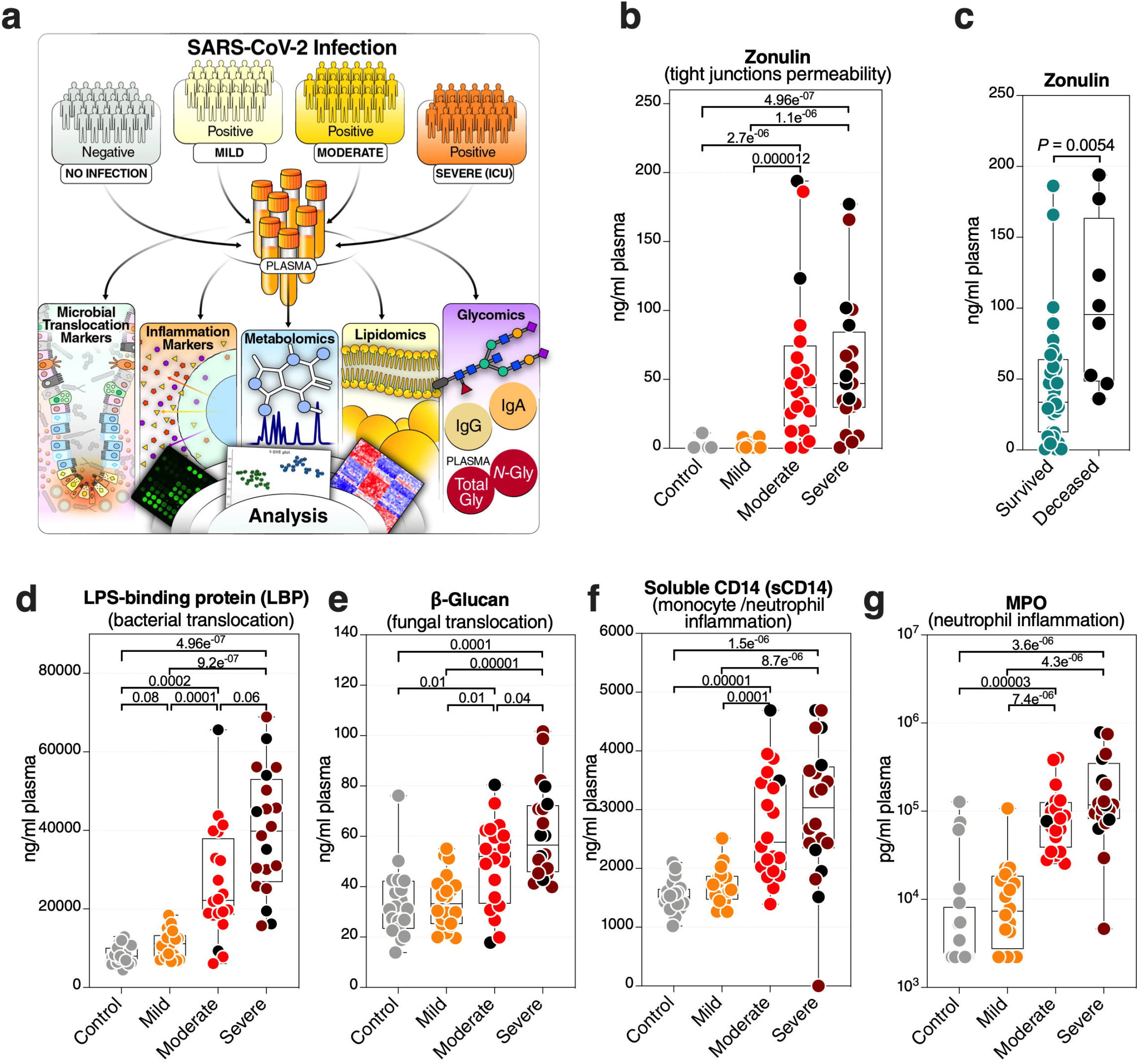
Severe COVID-19 is associated with an increase in markers of tight junction permeability and microbial translocation. **(a)** An overview of the main cohort study design; moderate and severe patients were hospitalized; severe indicates patients in the intensive care unit. **(b)** Levels of plasma zonulin, are higher during moderate and severe COVID-19 compared to mild COVID-19 or controls. Kruskal–Wallis test was used for statistical analysis. False discovery rate (FDR) was calculated using the Benjamini-Hochberg method. Symbols in black indicate deceased. **(c)** Zonulin levels are higher in hospitalized COVID patients (n=40) who eventually died from COVID-19 (n=8) compared to survivors (n=32). Nominal *P*-value was calculated using the Mann–Whitney U test. **(d-g)** Levels of LBP **(d)**, β-Glucan **(e)**, sCD14 **(f)**, and MPO **(g)**, are higher during severe COVID-19 compared to mild COVID-19 or controls. Kruskal–Wallis test was used for statistical analysis. FDR was calculated using Benjamini-Hochberg method. Black dots indicate deceased.

### Study validation cohort

Key measurements (zonulin, LBP, and soluble CD14) were confirmed using plasma samples from an independent cohort of 57 individuals tested positive for SARS-CoV-2 and 18 SARS-CoV-2 negative controls collected at RUMC. The 57 SARS-CoV-2 positives were selected to represent three disease states: 20 with mild symptoms (outpatients); 18 with moderate symptoms (inpatients hospitalized on regular wards); and 19 with severe symptoms (inpatients hospitalized in an ICU) (**Supplementary Table 2**).

### Ethics

All research protocols of the study were approved by the institutional review board (IRB) at Rush University (20070905-IRB01; approved July-27-2020) and at the Wistar Institute. All human experimentation was conducted in accordance with the guidelines of the US Department of Health and Human Services and those of the authors’ institutions.

### Measurement of plasma markers of tight junction permeability and microbial translocation

Plasma levels of soluble CD14 (sCD14), soluble CD163 (sCD163), LPS Binding Protein (LBP), and FABP2/I-FABP were quantified using DuoSet ELISA kits (R&D Systems; catalog # DY383-05, # DY1607-05, # DY870-05, and # DY3078, respectively). The plasma level of zonulin was measured using an ELISA kit from MyBiosorce (catalog # MBS706368). Levels of occludin were measured by ELISA (Biomatik; catalog # EKC34871). β-glucan detection in plasma was performed using Limulus Amebocyte Lysate (LAL) assay (Glucatell Kit, CapeCod; catalog # GT003). Plasma levels of Reg3A were measured by ELISA (RayBiotech; catalog # ELH-REG3A-1).

### Measurement of plasma markers of inflammation and immune activation

Plasma levels of GM-CSF, IFN-β, IFN-γ, IL-10, IL-13, IL-1β, IL-33, IL-4, IL-6, TNF-α, Fractalkine, IL-12p70, IL-2, IL-21, IL-22, IL-23, IP-10, MCP-2, MIP-1α, SDF-1a, IFN-α2a, IL-12/IL-23p40, and IL-15 were determined using customized MSD U□PLEX multiplex assay (Meso Scale Diagnostic catalog# K15067L-2). Plasma levels of C-Reactive Protein (CRP), Galectin-1, Galectin-3, and Galectin-9 were measured using DuoSet ELISA kits (R&D Systems; catalog # DY1707, # DY1152-05, # DY2045, and # DY1154, respectively). Levels of Growth Differentiation Factor-15 (GDF-15) were measured by ELISA using GDF-15 Quantikine ELISA Kit (R&D Systems; catalog # DGD150). Plasma levels of Myeloperoxidase (MPO), d-dimer, and C3a were measured by ELISA (Thermo Fischer; catalog # BMS2038INST, # EHDDIMER, #BMS2089, respectively).

### Untargeted measurement of plasma metabolites and lipids

Metabolomics analysis was performed as described previously (13). Lipidomics analysis was performed as described previously (14).

### IgG isolation

Bulk IgG was purified from plasma using Pierce Protein G Spin Plate (Thermo Fisher; catalog # 45204). IgG purity was confirmed by SDS gel.

### IgA isolation

Bulk IgA was purified from IgG depleted plasma using CaptureSelect IgA Affinity Matrix (Thermo Fisher; catalog # 194288010). IgA was concentrated using Amicon® filters (Milipore catalog #UFC805024), and purity was confirmed by SDS gel.

### N-glycan analysis using capillary electrophoresis

For both plasma and bulk IgG, *N*-glycans were released using peptide-N-glycosidase F (PNGase F) and labeled with 8-aminopyrene-1,3,6-trisulfonic acid (APTS) using the GlycanAssure APTS Kit (Thermo Fisher; catalog # A33952), following the manufacturer’s protocol. Labeled *N*-glycans were analyzed using the 3500 Genetic Analyzer capillary electrophoresis system. Total plasma *N*-glycans were separated into 24 peaks (**Supplementary Table 3**) and IgG *N*-glycans into 22 peaks (**Supplementary Table 4**). The relative abundance of *N*-glycan structures was quantified by calculating the area under the curve of each glycan structure divided by the total glycans using the Applied Biosystems GlycanAssure Data Analysis Software Version 2.0.

### Glycan analysis using lectin array

To profile plasma total and IgA glycomes, we used the lectin microarray as it enables analysis of multiple glycan structures. The lectin microarray employs a panel of 45 immobilized lectins with known glycan-binding specificity (lectins and their glycan-binding specificity are detailed in **Supplementary Table 5**). Plasma proteins or isolated IgA were labeled with Cy3 and hybridized to the lectin microarray. The resulting chips were scanned for fluorescence intensity on each lectin-coated spot using an evanescent-field fluorescence scanner GlycoStation Reader (GlycoTechnica Ltd.), and data were normalized using the global normalization method.

### Statistical analysis

Kruskal-Wallis and Mann–Whitney U tests were used for unpaired comparisons. Spearman’s rank correlations were used for bivariate correlation analyses. Severity correlation coefficient (SC *rho*) tested correlation versus patient groups with the severity groups quantified as follows: control=1, mild=2, moderate=3, severe=4. FDR for each type of comparison was calculated using the Benjamini–Hochberg approach within each data subset separately, and FDR<0.05 was used as a significance threshold. Principal Component Analysis was performed on log2-transformed z-scored data. Pathway enrichment analyses were done on features that passed significant SC *rho* at FDR<0.05. Enrichments for the metabolites were tested using QIAGEN’s Ingenuity® Pathway Analysis software (IPA®, QIAGEN Redwood City, www.qiagen.com/ingenuity) using the “Canonical Pathway” option. Enrichments for the lipids were done using LIPEA (https://lipea.biotec.tu-dresden.de/home) with default parameters. To explore biomarkers that could distinguish clinical outcome (hospitalization vs. non-hospitalization), a specific set of microbial translocation variables were identified among those with FDR<0.05. Variables for the multivariable logistic model were selected from the identified specific set of biomarkers using the Lasso technique with the cross-validation (CV) selection option by separating data in 5-fold. Due to the exploratory nature of this study with a moderate sample size, variable selection was determined using 100 independent rounds runs of CV Lasso with minimum tuning parameter lambda. The markers that were selected 80 or more times from 100 runs were used as a final set of variables in our model. The ability of the final logistic model was assessed by AUC with a 95% confidence interval. Statistical analyses were performed in R 4.0.2 and Prism 7.0 (GraphPad).

## RESULTS

### Characteristics of the study main cohort and study overview

We collected plasma samples from 60 individuals testing positive for SARS-CoV-2 (by RT-PCR) and 20 SARS-CoV-2 negative controls. The 60 SARS-CoV-2 positive individuals were selected to represent three disease states: 20 with mild symptoms (outpatients); 20 with moderate symptoms (inpatients hospitalized on regular wards); and 20 with severe symptoms (inpatients hospitalized in an intensive care unit (ICU)) (**Figure 1a**). Individuals were selected to have a median age between 52.5 to 58.5 years. There was no significant difference in age between groups (**Supplementary Figure 1**). The study cohort was also chosen to have a 35 to 60% representation of female gender per disease status group (**Supplementary Table 1**). Samples from hospitalized patients (moderate and severe groups) were collected at the time of diagnosis when the patient was admitted (**Supplementary Table 1**). Eight individuals of the cohort (two from the moderate group and six from the severe group) died from COVID-19 (**Supplementary Table 1**). The plasma samples from all individuals in this cohort were used in a multi-omic, systems biology approach that measured: markers of tight junction permeability and microbial translocation using ELISA and Limulus Amebocyte Lysate assays; inflammation and immune activation/dysfunction markers using ELISA and multiplex cytokine arrays; untargeted metabolomic and lipidomic analyses using mass spectrometry (MS); and plasma glycomes (from total plasma glycoproteins, isolated immunoglobulin G (IgG), and isolated immunoglobulin A (IgA)) using capillary electrophoresis and lectin microarray (**Figure 1a** and **Supplementary Table 6**).

### Severe COVID-19 is associated with high levels of markers of tight junction permeability and microbial translocation

We first asked whether severe COVID-19 is associated with differences in plasma markers of tight junction permeability and microbial translocation. We measured the plasma levels of eight established drivers and markers of intestinal barrier integrity (**Supplementary Table 6**). We found that severe COVID-19 is associated with high levels of zonulin (**Figure 1b**). Zonulin (haptoglobin 2 precursor) is an established mediator of tight junction permeability in the digestive tract, where higher levels of zonulin drive increases in tight junction permeability (15, 16). Notably, hospitalized individuals with higher plasma levels of zonulin were more likely to die compared to hospitalized individuals with lower levels of zonulin (**Figure 1c**).

These higher levels of zonulin could enable the translocation of microbes and their products from the gut into the blood, including parts of the cell wall of bacteria and fungus (17, 18). To test this supposition, we measured plasma levels of common bacterial and fungal markers. Exposure to bacterial endotoxin can be determined by measuring plasma lipopolysaccharide (LPS) binding protein (LBP). LBP is an acute-phase protein that binds to LPS to induce immune responses (19). Indeed, we observed high levels of LBP in individuals with severe COVID-19 compared to individuals with mild COVID-19 or controls (**Figure 1d)**. We also found higher levels of β-glucan, a polysaccharide cell wall component of most fungal species and a marker of fungal translocation (20), in individuals with severe COVID-19 compared to those with mild COVID-19 or controls (**Figure 1e**). In addition, there were significantly higher levels (FDR=0.025) of the tight junction protein occludin in the severe group compared to controls (data not shown). There also was a strong trend (FDR = 0.051) toward higher levels of the protein 3-alpha (REG3α), a marker of intestinal stress, comparing the severe and mild groups (data not shown). We did not observe high levels of intestinal fatty-acid binding protein (I-FABP), a marker of enterocyte apoptosis, suggesting that the high levels of tight junction permeability and microbial translocation are not associated with enterocyte death.

These high levels of tight junction permeability and microbial (both bacterial and fungal) translocation are expected to lead to microbial-mediated myeloid inflammation. Indeed, levels of soluble CD14 (sCD14; monocyte inflammation marker) (**Figure 1f**) and myeloperoxidase (MPO; neutrophil inflammation marker) (**Figure 1g**) were significantly higher during severe COVID-19 compared to mild and control groups. Levels of soluble CD163 (sCD163) were also higher significantly (FDR=0.04) in the severe group compared to controls (data not shown). These data suggest that COVID-19 severity and mortality are associated with plasma markers of higher tight junction permeability and higher translocation of bacterial and fungal products to the blood.

### Microbial translocation is linked to systemic inflammation

Higher levels of microbial translocation should lead to higher systemic inflammation. We measured the levels of 31 markers of systemic inflammation (**Supplementary Table 6**), including: 23 cytokines and chemokines (such as IL-6, IL-1β, MCP-1, IP-10, and TNFα), markers of inflammation and thrombogenesis (such as C-reactive protein (CRP) and D-dimer), a marker of complement activation (C3a), a marker of oxidative stress (GDF-15), and three immunomodulatory galectins (galectin-1, -3, and -9). As anticipated, the levels of many of these markers were higher in patients with severe COVID-19 compared to patients with mild COVID-19 or controls (**Figure 2a-left**). In particular, we observed higher levels of several cytokines and inflammatory markers. In addition to the expected changes, we also observed significantly higher levels of the immunomodulatory lectins, galectin-3 (**Figure 2b**) and galectin-9 (**Figure 2c**). Levels of Gal-9 were higher in the plasma of hospitalized patients who eventually died compared to survivors (**Figure 2d**). Last, notable dysregulations were observed in levels of C3a (**Figure 2e**; indicative of complement activation) and GDF-15 (**Figure 2f**; indicative of oxidative stress), with the levels of GDF-15 higher in deceased hospitalized patients compared to survivors (**Figure 2g**).

**Figure 2.**
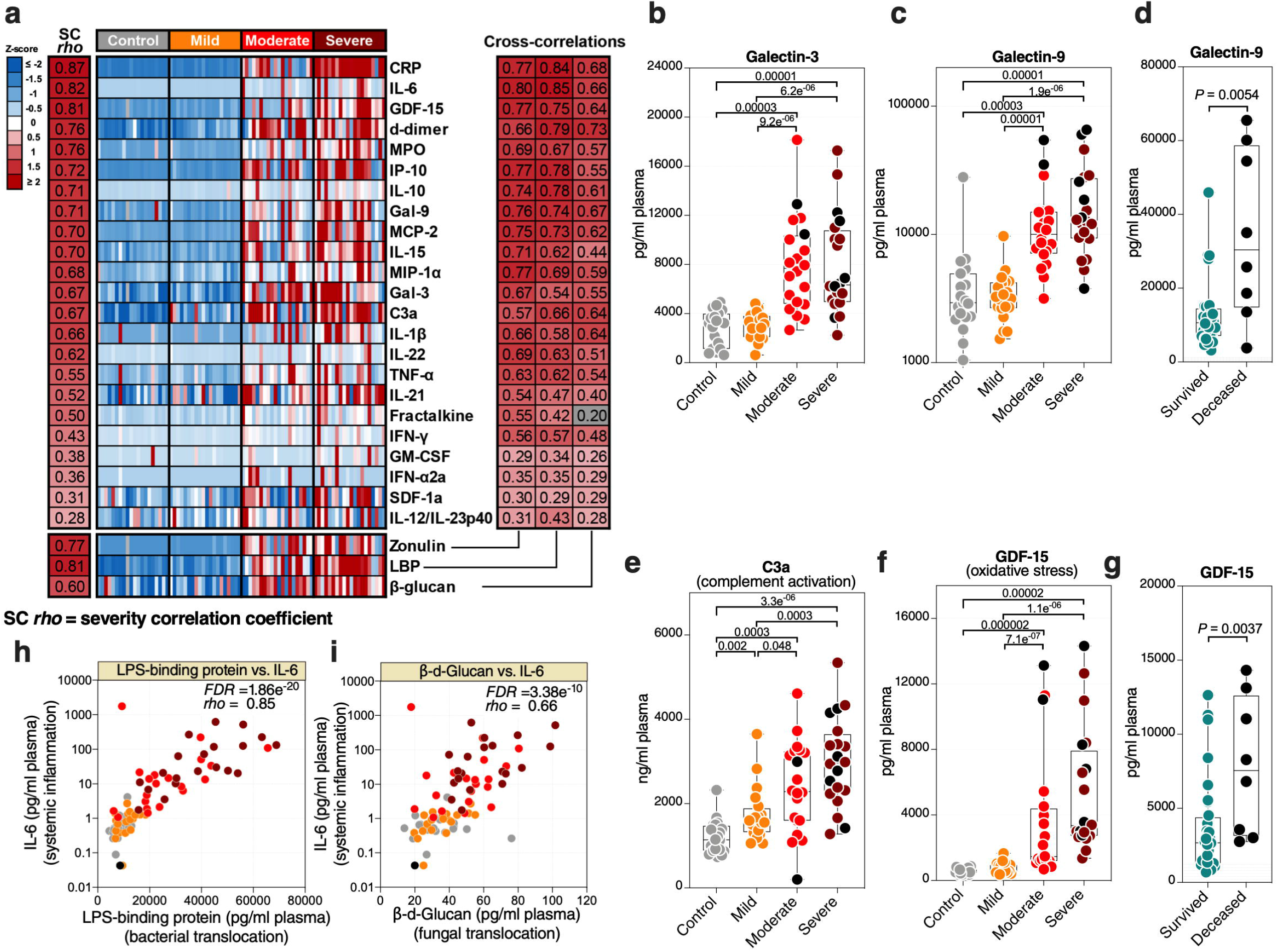
Markers of tight junction permeability and microbial translocation are associated with markers of systemic inflammation. **(a left)** Heat-map depicting plasma levels of 23 inflammation and immune activation/dysfunction markers whose levels are statistically (FDR<0.05) different between the four disease states. Statistical significance was determined using the Kruskal–Wallis test. FDR was calculated using Benjamini-Hochberg method. SC *rho* = coefficient of correlation with COVID-19 severity. Heat colors show standardized Z-scores across samples; red indicates upregulation, and blue indicates downregulation. **(a right)** Coefficients of correlation between zonulin, LBP, or β-Glucan and each of the 23 inflammation and immune activation/dysfunction markers. All red-colored correlations had statistical significance of FDR<0.05, whereas the grey-colored correlation was non-significant. Correlations were evaluated using Spearman’s rank correlation tests, and FDR was calculated using the Benjamini-Hochberg method. **(b-d)** Levels of representative variables, galectin-3 (Gal-3) **(b)** and galectin-9 (Gal-9) **(c)**, were higher during severe COVID-19 compared to mild COVID-19 or controls, with levels of Gal-9 higher among deceased hospitalized patients compared to survivors **(d). (e-g)** Levels of C3a **(e)** and GDF-15 **(f)** were higher during severe COVID-19 compared to mild COVID-19 or controls, with levels of GDF-15 higher among deceased hospitalized patients compared to survivors **(g)**. Kruskal–Wallis and Mann-Whitney tests were used for statistical analysis. FDR was calculated using Benjamini-Hochberg method. **(h-i)** Examples of correlations in (a) between LBP and IL-6 **(h)** or β-Glucan and IL-6 **(i)**. Spearman’s rank correlation tests were used for statistical analysis. Black dots indicate deceased.

Next, we examined the correlations between the markers of intestinal barrier integrity (zonulin) or microbial translocation (LBP and β-glucan) and the 31 markers of systemic inflammation and immune activation. As shown in **Figure 2a-right**, higher levels of zonulin, LBP, or β-glucan were strongly positively correlated with higher levels of many of the markers of systemic inflammation and immune activation, including IL-6 (**Figure 2h-i**). These data suggest that the potential disruption of the gut barrier integrity and microbial translocation during severe COVID-19 is associated with systemic inflammation. Our results do not imply that microbial translocation is the primary trigger of this inflammation, as it is likely that many pathophysiological pathways are involved in inflammation during COVID-19. However, the robust literature indicating that microbial translocation can fuel inflammation is consistent with our findings.

### Severe COVID-19 is associated with a plasma metabolomic profile that may reflect disrupted gut function

A second set of factors that may reflect the functional state of the gut and its microbiota are the plasma metabolites. Importantly, many of these are biologically active molecules that can directly impact immunological and inflammatory responses. We performed untargeted metabolomic analysis (using LC-MS/MS). Within the 80 plasma samples, we identified a total of polar 278 metabolites. We observed a significant metabolomic shift during severe COVID-19 (**Figure 3a**, a list of the top 50 dysregulated metabolites is in **Supplementary Figure 2**). Indeed, in principal component analysis of the full metabolomic dataset, the first component was able to completely distinguish controls (and mild patients) from those with severe disease. Pathway analysis of the COVID-19-dysregulated metabolites showed disruption in tRNA charging, citrulline metabolism, and several other amino acid (AA) metabolic pathways (**Figure 3b**, the top 10 dysregulated metabolic pathways are shown; **Supplementary Table 7** shows the top 50 dysregulated metabolic pathways with FDR<0.05). Importantly, changes in AA metabolism, including citrulline, arginine, methionine, and tryptophan (see **Figure 3b**) can influence the AA□metabolizing bacterial communities and disrupt the gut□microbiome immune axis (21). AA are absorbed and metabolized by enterocytes and gut microbiota. Consumption of AA by the gut microbiome is important for bacterial growth and is involved in the production of key microbiome-related metabolites (21).

**Figure 3.**
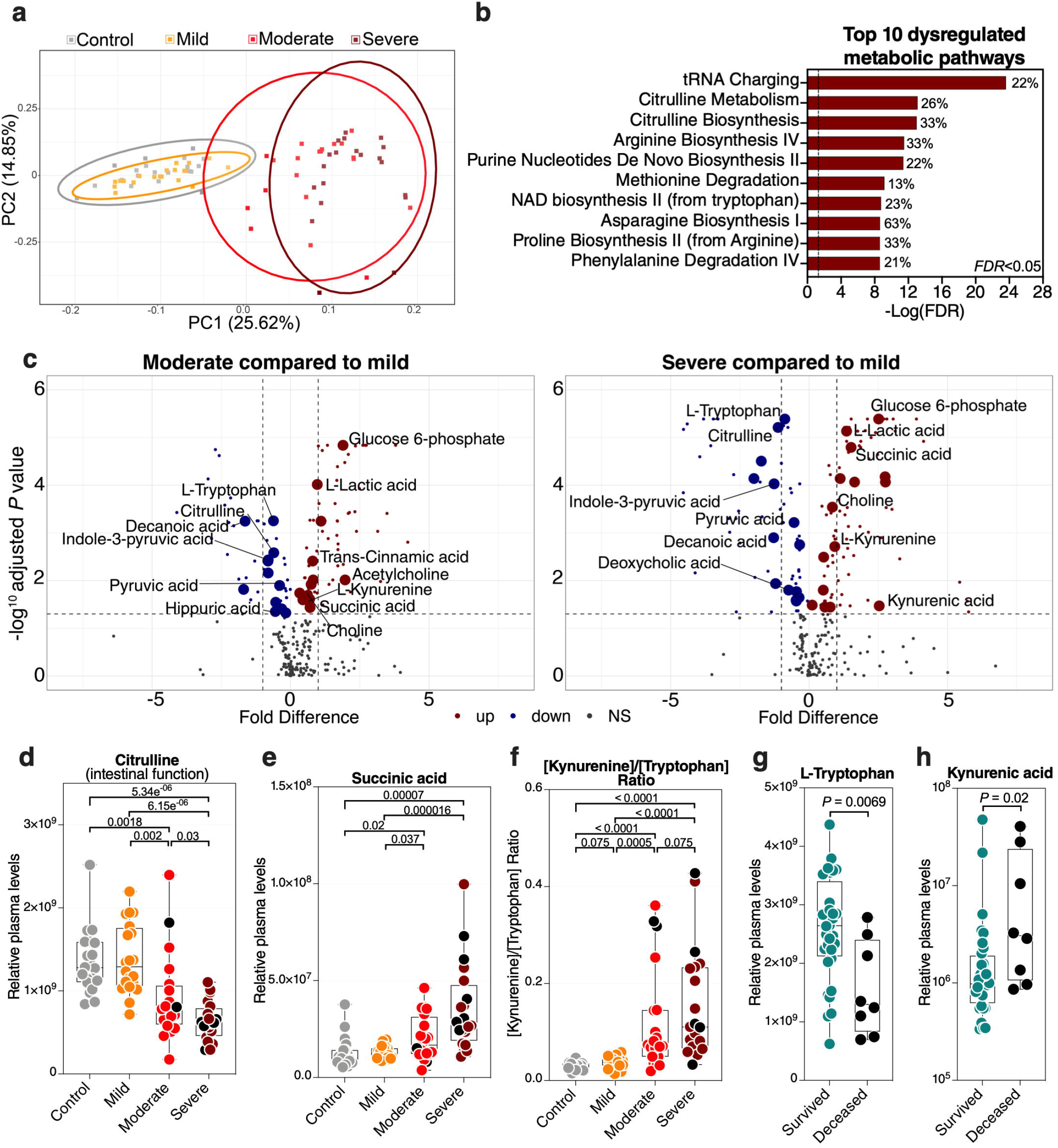
Severe COVID-19 is associated with metabolic dysregulation. **(a)** Principal component analysis (PCA) of the 278 metabolites identified in the plasma of the study cohort. Each symbol represents a study participant. **(b)** Ingenuity Pathway Analysis (IPA) of the plasma metabolites modulated between the disease states with FDR<0.05. The graph shows the top 10 dysregulated metabolic pathways with FDR<0.05. Percentages beside each pathway represent the ratio of dysregulated metabolites among the total number of metabolites assigned to this particular pathway in IPA. **(c)** Volcano plots depicting plasma metabolites dysregulated in the moderate group compared to the mild group (left) or the severe group compared to the mild group (right). NS= non-significant. The gut-associated metabolites (from Table 1) are indicated by the larger symbols, and a selected set is identified by name. **(d-f)** As representative examples, levels of citrulline are lower **(d)**, levels of succinic acid are higher **(e)**, and the ratio between kynurenine/tryptophan [Kyn/Trp] is higher **(f)** during severe COVID-19 compared to mild COVID-19 or controls. Kruskal–Wallis test was used for statistical analysis. FDR was calculated using Benjamini-Hochberg method. **(g-h)** For key metabolites in the tryptophan catabolism pathway, levels of tryptophan are lower **(g)**, and levels of kynurenic acid are higher **(h)** in deceased COVID-19 hospitalized patients compared to survivors. Nominal *P*-value was calculated using the Mann–Whitney U test. Black dots indicate deceased.

Next, we focused on 50 of the metabolites (out of the total of 278) that are known to be associated with the function of the gut and its microbiota (**Supplementary Table 8** lists the 50 metabolites and their references). Levels of most of these gut-associated plasma metabolites (35 out of 50) were dysregulated during severe COVID-19 compared to mild disease or controls (**Table 1** and **Figure 3c**). Within this metabolic signature of COVID-19-associated gut dysfunction is citrulline, which is also identified as a top metabolic pathway dysregulated by severe COVID-19 (**Figure 3b**). Citrulline is an amino acid produced only by enterocytes and an established marker of gut and enterocyte function (22). Its levels are significantly lower during severe COVID-19 (**Figure 3d)**. Also, within this metabolic signature is succinic acid, a marker of gut microbial dysbiosis, whose levels are higher during severe COVID-19 (**Figure 3e**).

Notable differences were also observed in several metabolites involved in the catabolism of the AA tryptophan (**Figure 3b, c**). Higher levels of tryptophan catabolism, indicated by high levels of kynurenine and low levels of tryptophan (i.e., the [Kyn/Trp] ratio), is a marker of gut microbial dysbiosis (23). Indeed, we observed a higher [Kyn/Trp] ratio in individuals with severe COVID-19 than in those with mild disease or controls (**Figure 3f**). Furthermore, lower levels of tryptophan and higher levels of kynurenic acid were associated with mortality among hospitalized COVID-19 patients (**Figure 3g-h**). Together, these data indicate that a metabolic signature associated with severe COVID-19 is compatible with disrupted gut functions and dysregulated gut□microbiome axis. However, it is important to note that many of these metabolic pathways are multi-faceted and can also reflect dysregulations in multiple-organ systems.

### Plasma metabolomic markers of COVID-19-associated gut dysfunction associate with higher inflammation and immune dysfunction

As noted above, many plasma metabolites are bioactive molecules that can directly impact immunological and inflammatory responses. Therefore, we sought to identify links between the 35 dysregulated gut-associated plasma metabolites (**Table 1**) and the dysregulated markers of microbial translocation, inflammation, and immune activation (**Figure 2a**). We observed strong links between levels of the dysregulated gut-associated metabolites and levels of markers of microbial translocation (**Figure 4a**) as well as levels of inflammation and immune activation (**Figure 4b**). Notable correlations were observed between lower levels of citrulline and higher IL-6 (**Figure 4c**), higher levels of succinic acid and higher IL-6 (**Figure 4d**), and higher [Kyn/Trp] ratio and higher IL-6 (**Figure 4e**). These data highlight the potential links between disrupted metabolic activities, especially those related to the gut and its microbiota, and systemic inflammation and immune dysfunction during COVID-19.

**Figure 4.**
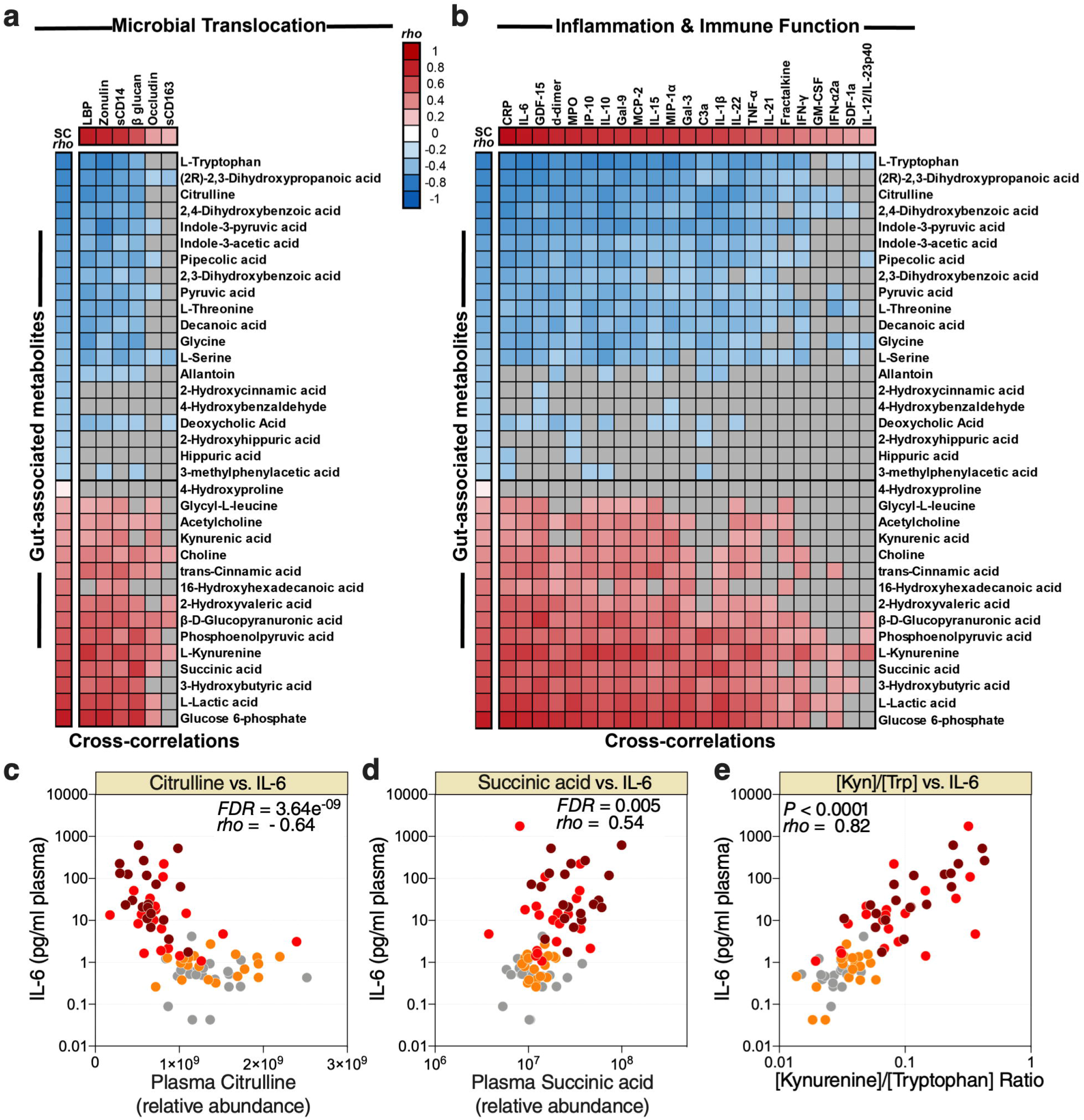
Metabolic markers of intestinal dysfunction are linked to microbial translocation and systemic inflammation. Correlation heat-maps depicting the correlations between COVID-19-modulated, gut-associated metabolites, and **(a)** markers of tight junction permeability and microbial translocation or (**b**) markers of inflammation and immune dysfunction. SC *rho* = coefficient of correlation with COVID-19 severity. Red-colored correlations = significant positive correlations with FDR<0.05, blue-colored correlations = significant negative correlations with FDR<0.05, gray-colored correlations = non-significant (FDR>0.05). **(c-e)** Examples of the correlations between citrulline and IL-6 **(c)**, succinic acid and IL-6 **(d)**, or [Kyn/Trp] ratio and IL-6 **(e)**. Spearman’s rank correlation tests were used for statistical analysis. FDR was calculated using Benjamini-Hochberg method.

### Severe COVID-19 is associated with disrupted lipid metabolism

Intermediary metabolites and sulfur-containing amino acids are potent modulators of lipid metabolism. Therefore, we performed lipidomic analysis and identified a total of 2015 lipids using untargeted MS. Similar to the plasma metabolome, the plasma lipidome shifted significantly during severe COVID-19 (**Figure 5a)**. These 2015 lipids were divided into 24 lipids classes (**Supplementary Table 9**); out of these 24 classes, 16 were significantly (FDR<0.05) different in the moderate and severe COVID-19 groups (11 were lower whereas five were higher compared to the mild or control groups) (**Figure 5b**). Pathway analysis of this severe-COVID-19-associated lipidomic signature showed that glycerophospholipid and choline metabolism were the most significantly dysregulated pathways (**Figure 5c**). The gut microbiota is heavily involved in these two interconnected pathways (24). Gut microbial dysbiosis can alter the digestion and absorption of glycerophospholipids, leading to several diseases (25, 26).

**Figure 5.**
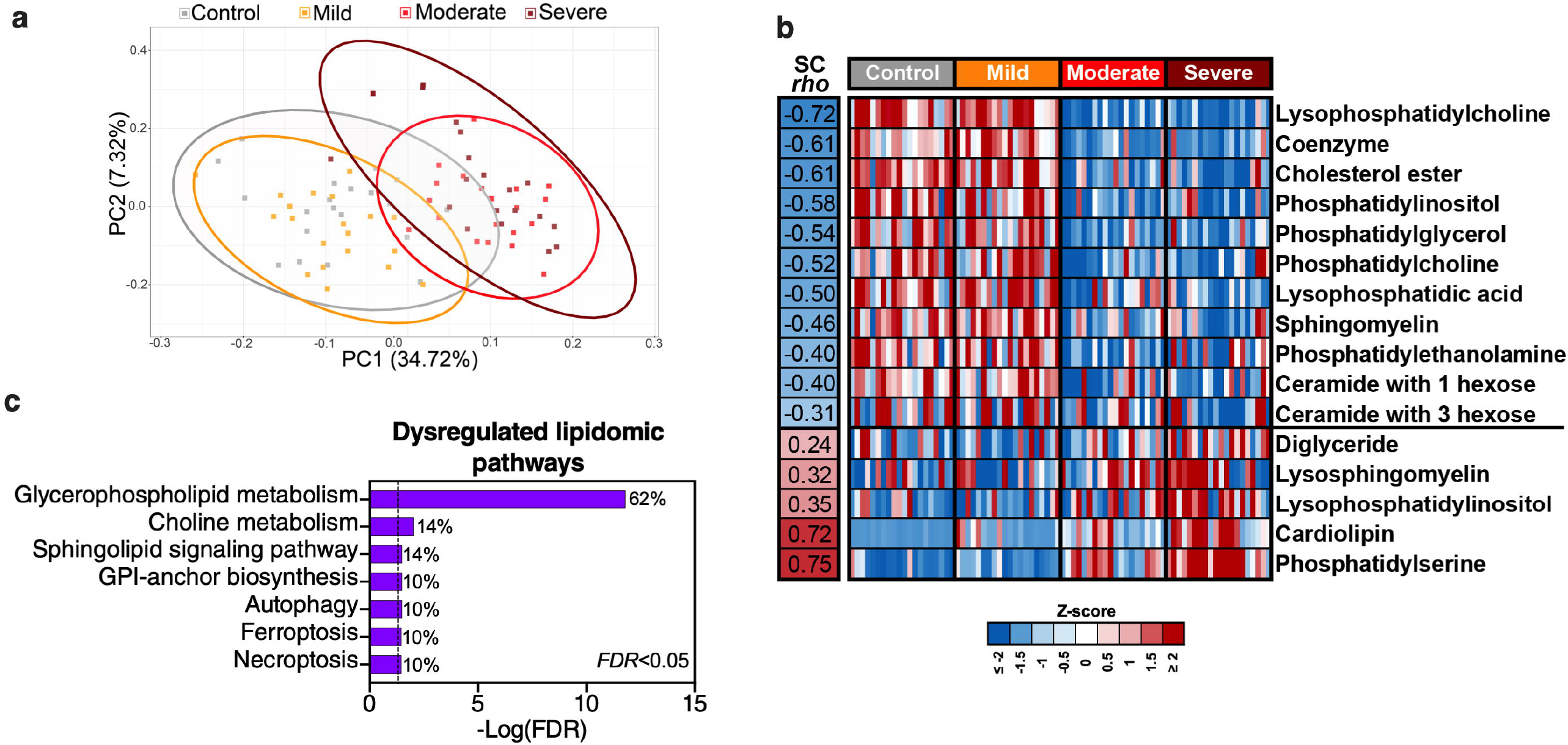
Severe COVID-19 is associated with disrupted lipid metabolism. **(a)** Principal component analysis (PCA) of 2015 lipids identified in the plasma of the study cohort. **(b)** The 2015 identified lipids were assigned to 24 classes (Supplementary Table 5). Heat-map depicts the 16 lipid classes dysregulated by severe COVID-19 (FDR<0.05). Statistical significance was determined using the Kruskal–Wallis test. FDR was calculated using Benjamini-Hochberg method. SC *rho* = coefficient of correlation with COVID-19 severity. Heat colors show standardized Z-scores across samples; red indicates upregulation, and blue indicates downregulation. **(c)** Lipid pathway analysis of the plasma lipids modulated between the disease states with FDR<0.05 was performed using LIPEA (Lipid Pathway Enrichment Analysis; https://lipea.biotec.tu-dresden.de/home). The graph includes all dysregulated pathways with FDR<0.05. Percentages beside each pathway represent the ratio of dysregulated lipids among the total number of lipids assigned to this particular pathway by LIPEA.

It is also known that COVID-19 severity is linked to pre-existing cardiometabolic-associated diseases (27). Furthermore, COVID-19 itself can cause liver dysfunction (28). Indeed, many of the individuals in our main cohort with moderate and severe COVID-19 had diabetes and/or high blood pressure. We sought to examine whether these conditions contribute to our main findings. We examined the differences in the levels of zonulin, LBP, β-glucan, sCD14, and IL-6 between hospitalized patients (moderate and severe groups) who had diabetes or not (**supplementary Figure 3a**), or patients who had high blood pressure or not (**supplementary Figure 3b**). We did not observe any significant difference in the levels of these selected markers between these groups. However, the contribution of pre-existing metabolic conditions and post-infection intestinal and liver complications to the observed disrupted plasma profiles warrant further investigation.

### Severe COVID-19 is associated with altered plasma glycomes that are linked to inflammation and complement activation

Finally, we examined plasma glycomes. It has been reported that translocation of glycan-degrading enzymes released by several members of the gut microbiome can alter circulating glycomes (10). Within the plasma glycome, glycans on circulating glycoproteins and antibodies (IgGs and IgAs) play essential roles in regulating several immunological responses, including complement activation (9). For example, galactosylated glycans link Dectin-1 to Fcγ receptor IIB (FcγRIIB) on the surface of myeloid cells to prevent inflammation-mediated by complement activation (9). A loss of galactose decreases the opportunity to activate this anti-inflammatory checkpoint, thus promoting inflammation and complement activation, including during IBD (10). Indeed, IgG glycomic alterations associate with IBD disease progression, and IBD patients have lower IgG galactosylation compared to healthy controls (11).

We applied several glycomic technologies to analyze the plasma glycome (total plasma, isolated IgG, and isolated IgA). First, we used capillary electrophoresis to identify the *N*-linked glycans of total plasma glycoproteins and isolated plasma IgG (this identified 24 and 22 glycan structures, respectively; their names and structures are in **Supplementary Tables 3 and 4**). We also used a 45-plex lectin microarray to identify other glycans on total plasma glycoproteins and isolated IgA. The lectin microarray enables sensitive analysis of multiple glycan structures by employing a panel of 45 immobilized lectins (glycan-binding proteins) with known glycan-binding specificity (**Supplementary Table 5** lists the 45 lectins and their glycan-binding specificities) (29).

We first observed significant (FDR<0.05) glycomic differences during severe COVID-19 in levels of IgA glycans, plasma *N*-glycans, plasma total glycans, and IgG glycans (**Figure 6a)**. These changes are exemplified by an apparent loss of the anti-complement activation galactosylated glycans from IgG and total plasma glycoproteins (**Figure 6b-c**, respectively). When we examined the correlations between the plasma glycome and markers of tight junction permeability/microbial translocation or inflammation/immune activation (**Supplementary Figure 4**), as expected, we observed significant negative correlations (FDR<0.05) between levels of terminal galactose on IgG or plasma glycoproteins and markers of permeability/ translocation (**Figure 6d**) or markers of inflammation (**Figure 6e**). These data highlight the potential links between the disrupted plasma glycome and systemic inflammation during COVID-19.

**Figure 6.**
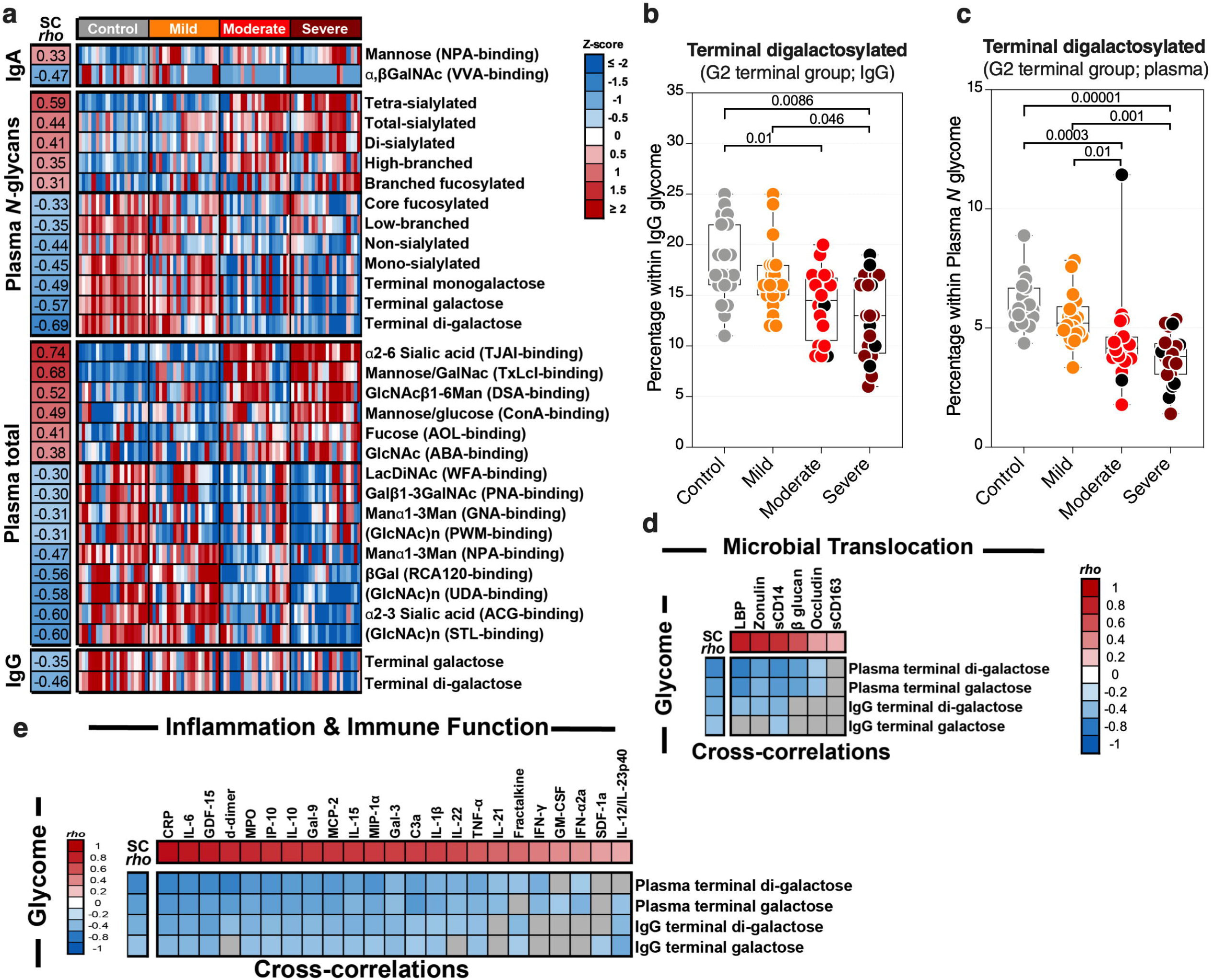
Severe COVID-19 is associated with plasma glycomic dysregulations. **(a)** Heat-map depicting glycans dysregulated by severe COVID-19 (FDR<0.05). Names of the glycan-binding lectins are provided in parentheses. Statistical significance was determined using the Kruskal–Wallis test. FDR was calculated using Benjamini-Hochberg method. SC *rho* = coefficient of correlation with COVID-19 severity. Heat colors show standardized Z-scores across samples; red indicates upregulation, and blue indicates downregulation. **(b-c)** Levels of terminal digalactosylated *N*-glycans in IgG **(b)** or total plasma glycoproteins **(c)** are lower during severe COVID-19 compared to mild COVID-19 or controls. Kruskal–Wallis test. FDR was calculated using Benjamini-Hochberg method. **(e-d)** Correlation heat-maps depicting the correlations between galactosylated *N*-glycans (rows) and markers of tight junction permeability and microbial translocation **(d)** or markers of inflammation and immune dysfunction (**e**). SC *rho* 552 = coefficient of correlation with COVID-19 severity. Red-colored correlations = significant positive correlations with FDR<0.05, blue-colored correlations = significant negative correlations with FDR<0.05, and gray-colored correlations = non-significant. Spearman’s rank correlation tests were used for statistical analysis. FDR was calculated using Benjamini-Hochberg method. Black dots indicate deceased.

### Multivariable logistic models, using cross-validation Lasso technique, selected gut-associated variables whose combination associates with the risk of hospitalization during COVID-19

Our data thus far support the hypothesis that gut dysfunction may contribute to COVID-19 severity. We sought to examine whether markers of tight junction permeability and microbial translocation (**Supplementary Table 6**) can distinguish between hospitalized COVID-19 patients (moderate and severe groups combined) and non-hospitalized individuals (mild and controls combined). We applied the machine learning algorithm Lasso (least absolute shrinkage and selection operator) regularization to select markers with the highest ability to distinguish between the two groups. The analysis employed samples with complete data sets (n=79; one sample did not have complete data). Lasso selected zonulin, LBP, and sCD14 as the three markers to be included in a multivariable logistic regression model that distinguishes hospitalized from non-hospitalized individuals with area under the ROC curve (AUC) of 99.23% (**Figure 7a-b**; 95% confidence interval: 98.1% -100%). This value was higher than the AUC values obtained from logistic models using each variable individually (**Table 2)**. Next, we used the multivariable logistic model to estimate a risk score of hospitalization for each individual. We then examined the ability of these risk scores to classify hospitalized from non-hospitalized individuals. As shown in **Figure 7c**, the model correctly classified 97.5% of hospitalized (sensitivity) and 94.9% of non-hospitalized (specificity) individuals, with an overall accuracy of 96.2%. Furthermore, we examined the ability of the L-kynurenine/L-tryptophan [Kyn/Trp] ratio, an established marker of gut microbial dysbiosis described above, to distinguish hospitalized from non-hospitalized individuals. Logistic model showed that [Kyn/Trp] ratio alone can distinguish hospitalized from non-hospitalized with an AUC value of 91.9% (**Figure 7d**; 95% confidence interval: >85% -98.7%). These data raise the possibility that some of these markers may be able to predict the risk of disease progression if measured immediately after diagnosis. Markers of intestinal barrier permeability have been used as predictors of multiple organ dysfunction during critical illness (30). Future longitudinal, controlled studies will be needed to assess this possibility.

**Figure 7.**
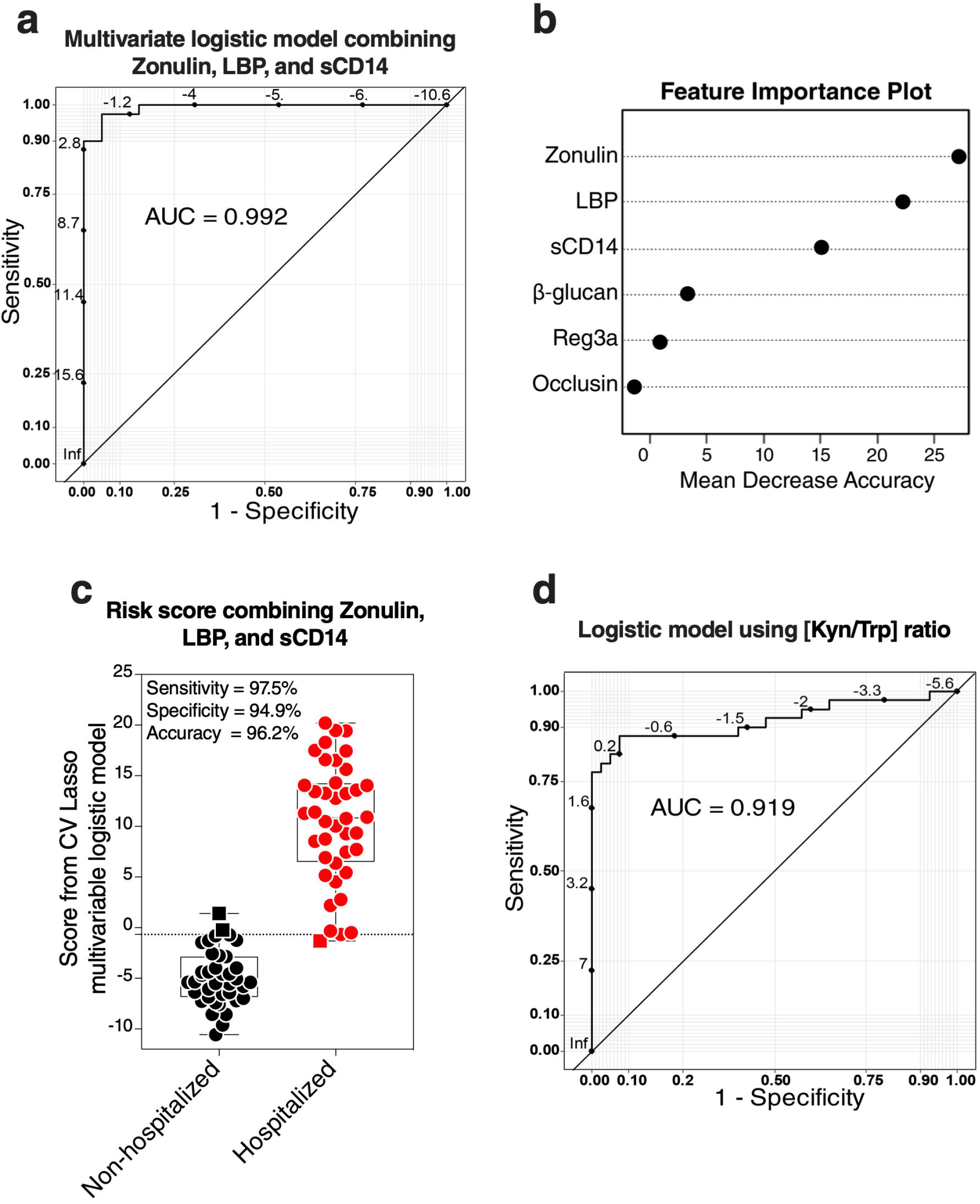
Logistic models using markers of tight-junction permeability and microbial translocation distinguish hospitalized from non-hospitalized individuals. **(a-b)** The machine learning algorithm, Lasso (least absolute shrinkage and selection operator) regularization, selected three markers (zonulin, LBP, and sCD14) that, when combined, can distinguish hospitalized from non-hospitalized individuals. (a) The receiver operator characteristic (ROC) curve from the multivariable logistic regression model with the three variables combined. (b) feature importance plot. (**c**) Coefficients from the multivariable logistic model were used to estimate a hospitalization risk score for each individual and then tested for the ability of these scores to accurately classify hospitalized (n=40) from non-hospitalized (n=39; one sample did not have a complete dataset) individuals at an optimal cut-point. Squares represent individuals the model failed to identify correctly. **(d)** Logistic regression model using the L-kynurenine/L-tryptophan [Kyn/Trp] ratio is able to distinguish hospitalized from non-hospitalized individuals. ROC curve showing the area under the curve (AUC) is 91.3%.

### Zonulin, LBP, and sCD14 plasma levels are higher during severe COVID-19 in an independent validation cohort

Finally, we sought to confirm some of our key findings in an independent cohort of 57 individuals tested positive for SARS-CoV-2 and 18 SARS-CoV2 negative controls. The 57 SARS-CoV-2 positives were selected to represent three disease states: 20 with mild symptoms (outpatients); 18 with moderate symptoms (inpatients hospitalized on regular wards); and 19 with severe symptoms (inpatients hospitalized in an ICU) (**Supplementary Table 2**). We focused on three measurements, zonulin, LBP, and sCD14, as these three measurements together were able to distinguish hospitalized from non-hospitalized individuals in the main cohort (Figure 7a-c). We observed higher levels of zonulin, LBP, and sCD14 during severe COVID-19 in this validation cohort (**Figure 8a-c**). Furthermore, we validated our multivariable logistic model in Figure 7a-c using data from this validation cohort. A combination of zonulin, LBP, and sCD14 was able to distinguishe hospitalized from non-hospitalized individuals in the validation cohort with AUC of 88.6% (95% confidence interval: 80.3% -96.8%; **Table 3**). This analysis further highlights the plausible link between severe COVID-19 and disrupted gut function.

**Figure 8.**
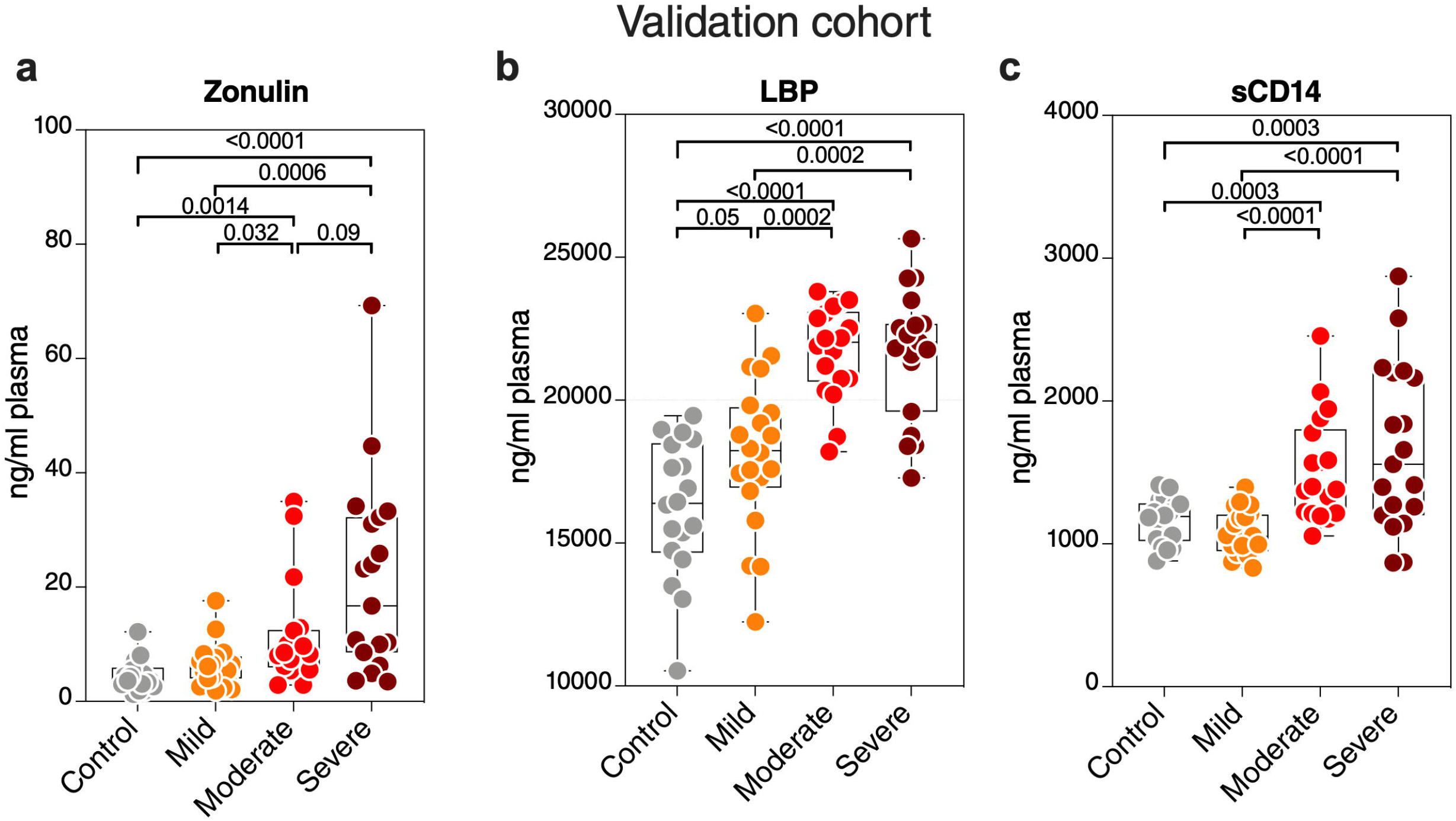
Validation of key measurements in an independent cohort. Levels of plasma **(a)** zonulin, **(b)** LBP, and **(c)** sCD14 are higher during moderate and severe COVID-19 compared to mild COVID-19 or controls in an independent validation cohort. Kruskal–Wallis test as used for statistical analysis.

## DISCUSSION

We used a systems biology approach to provide multiple layers of evidence that severe COVID-19 is associated with markers of disrupted intestinal barrier integrity, microbial translocation, and intestinal dysfunction. These data highlight disruption in gut barrier integrity as a potential force that may contribute to COVID-19 severity. Our data are compatible with previous reports showed that severe COVID-19 is associated with bacterial translocation to the blood and increased levels of microbial-associated immune activation markers (31, 32). Our results do not imply that microbial dysbiosis and translocation are the primary triggers of severe COVID-19, as the complex clinical syndrome of severe COVID-19 likely embodies multiple pathophysiological pathways. Also, our *in vivo* analyses do not unequivocally demonstrate a causal relationship between gut dysfunction and COVID-19 severity. However, the robust literature indicating that a disrupted intestinal barrier and microbial dysbiosis and translocation fuel inflammation and disease severity during ARDS (3, 4) supports our hypothesis and is consistent with our findings.

SARS-CoV-2 infection can affect the gastrointestinal tract (GI) tract and cause GI symptoms (33). Recently, it has been suggested that the severity of GI symptoms (mainly vomiting and diarrhea) correlates inversely with COVID-19 severity (for unclear reasons) (34). On the other hand, our observations suggest that disruption in gut function and higher microbial translocation correlate positively with COVID-19 severity. These are not necessarily mutually exclusive findings, but rather indicate that the interplay between the gut and SARS-CoV2 infection in modulating disease severity is complex. The potential role of the gut should be further explored, in multiple cohorts and settings, longitudinally during different stages of infection, and using gut biopsies and stool samples. Also, it will be important to examine, in future studies, the location and extent of these potential barrier defects along the length of small and large intestine, using gut biopsies from infected patients and controls.

Intestinal disruption during SARS-CoV-2 infection could be caused directly, and/or indirectly. Directly, SARS-CoV-2 can infect gut cells (5); other viral infections of the gut can change gut structure and cause breakdown of the epithelial barrier (6). Indirectly, lung infection or injury leads to systemic inflammation (including a cytokine storm), which then disrupts gut barrier integrity, increasing the permeability to gut microbes and microbial products. Whether the potential disruption of the gut barrier during severe COVID-19 is caused directly by infection and/or indirectly by systemic inflammation and cytokines is not known but warrant further investigations. Also, it will be important to examine the impact of this potential microbial translocation on immune cell functions both in the intestines and systemically.

Our data raise several critical questions, including are there long-term implications of the potential disrupted gut barrier and intestinal function in survivors of severe COVID-19? In survivors of SARS-CoV-1 infection, long-term health complications (including metabolic dysfunctions) were observed for many years after convalescence (35). HIV+ individuals also can suffer complications of gut microbial translocation for years after viral suppression (36) The current ‘long-haulers’ (37, 38) after severe COVID-19 may also be on a path towards long term consequences due to persistent, microbial translocation. Understanding the long-term implications of the potential disrupted gut function during severe COVID-19 should be a clinical priority. An accompanying priority should be to consider how to modify clinical practice to prevent or reduce gut disruption. Currently, for example, a large number of patients are receiving antibiotic therapy during their COVID-19 treatment (39). However, massive use of antibiotics can alter gut microbiota and gut function, leading to higher susceptibility to inflammatory disorders (40). Thus, for any clinical practices that alter the gut, their overall impact on disease course should be carefully considered.

Our study also reveals several potential therapeutic targets for severe COVID-19, including zonulin. Zonulin is an established modulator of the intestinal tight junctions (15). Microbial dysbiosis and translocation enhance zonulin release, which in turn induces tight junction permeability, leading to more microbial translocation. This microbial translocation triggers inflammation, which promotes further gut leakiness (17, 18). Increased intestinal permeability and serum zonulin levels have been observed during many inflammatory diseases, including Crohn’s disease (41). Preventing zonulin-mediated increase in intestinal permeability by a zonulin receptor antagonist AT1001 (larazotide acetate) decreased the severity and incidence of several inflammation-associated diseases in pre-clinical and clinical studies (42-44). The high levels of serum zonulin we observed during severe COVID-19, which were associated with inflammation and mortality, raise the question of whether modulators of tight junction permeability (such as with AT-1001) can lessen COVID-19 severity.

Our data show a disruption in several multi-faceted metabolic pathways, some of which are linked to gut functions. Plasma citrulline levels were lower in both moderate and severe COVID-19 patients compared to the mild and control groups, and the citrulline metabolism and biosynthesis pathways were among the top metabolic pathways disrupted in severe COVID-19. Citrulline is an intermediate in arginine metabolism (45), and is an established marker of gut and enterocyte function (22, 46). Disrupted citrulline metabolism, as we observed during severe COVID-19, has been associated with microbial dysbiosis and dysregulated intestinal function (47). However, ACE2 blocking is an alternative explanation of the low levels of citrulline. ACE2 is required for the function of amino acid transporters (48). Thus, the binding of SARS-CoV-2 to ACE2 may reduce the functions of amino acid transporters, leading to the reduction of citrulline and other amino acids such as tryptophan. Additionally, plasma citrulline levels might be an indication of defects in liver and/or kidney functions (49). We also observed high levels of succinic acid and kynurenic acid. Both metabolites have been associated with intestinal microbial dysbiosis (23, 50). However, succinic acid has also been associated with mucosal hypoxia (50). To what extend the disruption of intestinal barrier integrity and microbial dysbiosis contribute to these disrupted metabolic pathways should the subject of future studies. These studies will help us to better understand the interaction between the gut, intestinal microbiota, and amino acid metabolism during COVID-19. This understanding might be critical to inform pharmaceutical and diet approaches to improve COVID-19 outcomes.

A significant strength of our multi-omics approach is its ability to uncover connections between severe COVID-19 and biomolecules of different classes. The carbohydrate structures (glycans) attached to circulating proteins, including antibodies, and their receptors (lectins) are increasingly being appreciated for their essential roles in a variety of immune functions. Among the glycobiological molecules regulated by severe COVID-19 are galectins (increasing) and galactosylated glycans on circulating glycoproteins (decreasing). Both may point to potential glycomic contributions to the severity of COVID-19. First, galectins (secreted, GalNAc-binding proteins) have emerged as significant modulators of cytokine expression by immune cells during several diseases, including viral infections (51). Importantly, small molecule inhibitors for galectins, especially for Gal-3, can reduce inflammation and cytokine release (52). Therefore, galectins represent potential therapeutic targets to reduce cytokine storm during COVID-19 (53).

Caveats of our study include the sample size and sampling of blood. As noted above, correcting for potential confounders (such as ethnicity) will require larger cohorts from varying geographic and demographic settings. Additional independent test sets and samples from non-COVID1-9 hospitalized and ICU-admitted patients will also be needed. Longitudinal analyses will be required to examine the long-term implications of our findings and their potential value as prognostic biomarkers. Analysis of gut biopsies, stool, and bronchial lavage will be needed to determine the precise contributions of the gut-lung axis in COVID-19. Analyzing the impact of the potential microbial translocation during severe COVID-19 on particular immune cell (such as Th17) in the intestines and systemically, will be also needed. Furthermore, environmental differences in diet and social environment may impact intestinal inflammation, therefore, future studies to understand the potential impact of socioeconomic status on the potential link between microbial translocation and severe COVID-19 will be needed. Finally, mechanistic studies *in vitro* and in animal models of SARS-CoV2 infection will be needed to examine the direct versus the indirect impact of the infection on intestinal barrier integrity and function.

In summary, while this study was exploratory in nature, our data suggest: (1) severe COVID-19 is associated with markers of a) disrupted intestinal barrier integrity; b) higher microbial translocation; and c) gut dysfunction; (2) severe COVID-19 is associated with a dramatic shift in levels of several biologically active molecules, which likely contribute to disease severity by inducing inflammation. Our study is beginning to shed light on the potentially critical role of intestinal barrier integrity in the pathophysiology of severe COVID-19. By understanding these underappreciated underpinnings of COVID-19, this work may serve to identify biomarkers for risk stratification and build a foundation for developing strategies to prevent or reduce the severity of COVID-19.

## Data Availability

All data are available in the manuscript or upon request.

## TABLES

**Table 1**. Levels of the 35 (out of 50) gut- and gut microbiota-associated plasma metabolites that are disrupted during COVID-19. Red indicates upregulation, blue indicates downregulation; color intensity indicates larger difference. Green indicates FDR<0.05; color intensity indicates lower FDR.

**Table 2**. Results of logistic regression models of tight junction permeability and microbial translocation markers for ability to distinguish hospitalized from non-hospitalized individuals.

**Table 3**. AUC values of the logistic regression model built with data from the main cohort and validated with data from the validation cohort.

## SUPPLEMENTARY MATERIALS

**Supplementary Figure 1: Age was not significantly different between the groups in the main cohort**. Kruskal–Wallis test was used for statistical analysis

**Supplementary Figure 2: Top 50 metabolites dysregulated by COVID-19**. Heat-maps depicting the top 25 metabolites induced and the top 25 metabolites reduced by COVID-19 (FDR<0.05). Statistical significance was determined using the Kruskal–Wallis test. FDR was calculated using Benjamini-Hochberg method. SC *rho* = coefficient of correlation with COVID-19 severity. Heat colors show standardized Z-scores across samples; red indicates upregulation, and blue indicates downregulation.

**Supplementary Figure 3: Levels of key markers are not different between groups with different diabetes or blood pressure status. (a)** No = non-diabetic; Pre = Pre-diabetic; and Yes = diabetic. Statistical significance was determined using the Kruskal–Wallis test (ns = non-significant; *P*>0.05). **(b)** No = not suffering from high blood pressure and Yes = suffering from high blood pressure. Statistical significance was determined using the Mann–Whitney U test (ns = non-significant; *P*>0.05).

**Supplementary Figure 4: The COVID-19-associated plasma glycomic signatures are associated with microbial translocation and systemic inflammation**. Correlation heat-maps depicting the correlations between COVID-19 modulated plasma glycans and **(a)** markers of tight junction permeability and microbial translocation or (**b**) markers of inflammation and immune dysfunction. SC *rho* = coefficient of correlation with COVID-19 severity. Red-colored correlations = significant positive correlations with FDR<0.05, blue-colored correlations = significant negative correlations with FDR<0.05, and gray-colored correlations = non-significant correlations.

**Supplementary Table 1**. Demographic and clinical characteristics of the study cohort.

**Supplementary Table 2**. Demographic characteristics of the validation cohort.

**Supplementary Table 3**. The structures and names of *N*-glycans identified in plasma by capillary electrophoresis. These glycan structures can be grouped into 15 groups: bisecting GlcNAc (B group), sialic acid (non-sialylated (S0), mono-sialylated (S1), di-sialylated (S2), tri-sialylated and (S3), tetra-sialylated (S4), and total sialylated (ST)), galactose (agalactosylated (G0), mono-galactosylated (G1), di-galactosylated (G2), and total galactosylated (GT)), core fucose (FC group), branched fucose (FB group), high branched (HB group), and low branch (LB group).

**Supplementary Table 4**. The structures and names of *N*-glycans identified in IgG by capillary electrophoresis. These glycan structures were grouped into 9 groups, depending on the presence or absence of four key monosaccharides: bisecting GlcNAc (B group), sialic acid (mon-sialylated (S1), di-sialylated (S2), and total sialylated (ST)), terminal galactose (agalactosylated (G0), mono-galactosylated (G1), di-galactosylated (G2), and total galactose (GT)), and fucose (F group).

**Supplementary Table 5**. Lectins used in the 45-plex lectin microarray and their glycan-binding specificity.

**Supplementary Table 6:** A list of plasma markers measured in this study.

**Supplementary Table 7:** Top 50 metabolic pathways disrupted by severe COVID-19.

**Supplementary Table 8:** List of the gut-associated and gut microbiota-associated metabolites detected in our study using untargeted LC-MS/MS (50 of the 278 metabolites identified in plasma).

**Supplementary Table 9**. The two thousand fifteen lipids identified in this study were assigned to 24 lipid classes.

## ACKNOWLEDGMENTS

This study is supported by a supplement to the NIH R01 DK123733 (R01 DK123733-01S1) for M.A-M, A.L, and A.Ke and R24 AA026801-02S1 for A.Ke. M.A-M is also supported by The Foundation for AIDS Research (amfAR) impact grant # 109840-65-RGR, NIH grants (R01 AG062383, R01NS117458, R21 AI143385, R21 AI129636, and R21 NS106970), and the Penn Center for AIDS Research (P30 AI 045008). The Wistar Proteomics and Metabolomics Shared Resource is supported in part by NIH Cancer Center Support Grant CA010815. The Thermo Q-Exactive HF-X mass spectrometer was purchased with NIH grant S10 OD023586. We would like to thank Rachel E. Locke, Ph.D., for providing comments.

## AUTHOR CONTRIBUTIONS

M.A-M, A.L, and A.Ke conceived and designed the study. L.B.G. carried out the majority of experiments. H.D., X.Y, H.W, Q.L, and A.Ko performed all bioinformatics and biostatistical analyses. M.D. ran the lectin array experiments. C.S.P reviewed and selected gut-associated metabolites. N.F.Z, R.A.B, M.W.S, CBF, A.Ke, and A.L selected study participants and interpreted data. A.R.G, N.G, and H.T performed metabolic and lipidomic analyses. L.B.G. and M.A-M wrote the manuscript, and all authors edited it.

## COMPETING INTERESTS STATEMENT

Authors have no competing interests.

## Notes

### Competing Interest Statement

The authors have declared no competing interest.

### Author Declarations

Ethics. All research protocols of the study were approved by the institutional review board (IRB) at Rush University (20070905-IRB01; approved July-27-2020). All human experimentation was conducted in accordance with the guidelines of the US Department of Health and Human Services and those of the authors institutions.

