## Supplementary figures and images for "Plasma Markers of Disrupted Gut Permeability in Severe COVID-19 Patients"

### Supplementary_Figure_1

Supplementary Figure 1

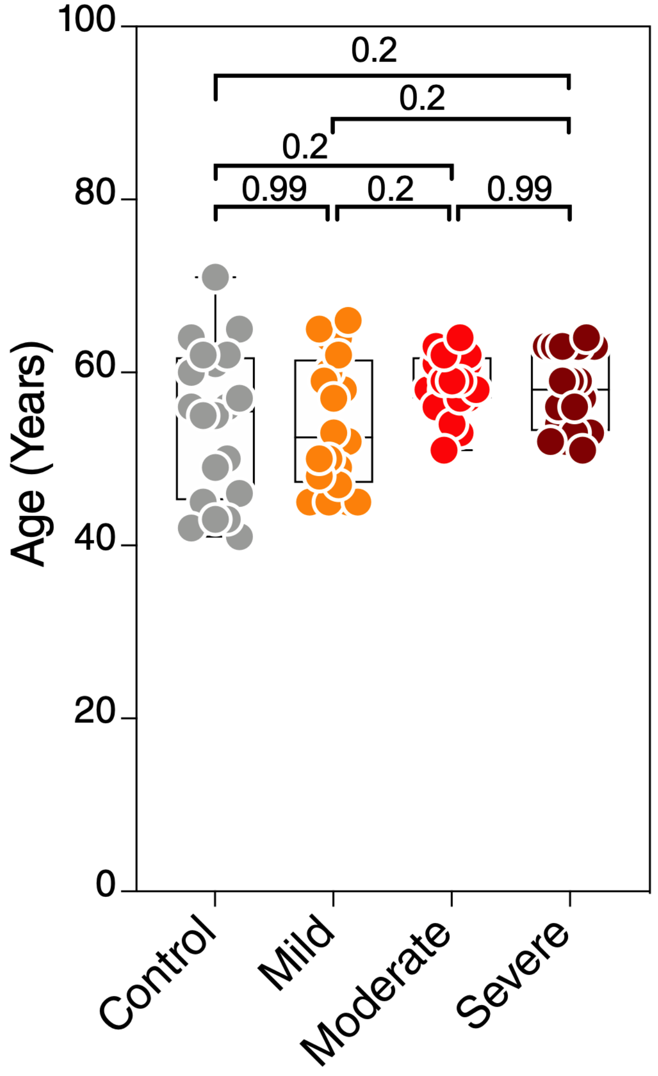

### Supplementary_Figure_2

#### Supplementary Figure 2

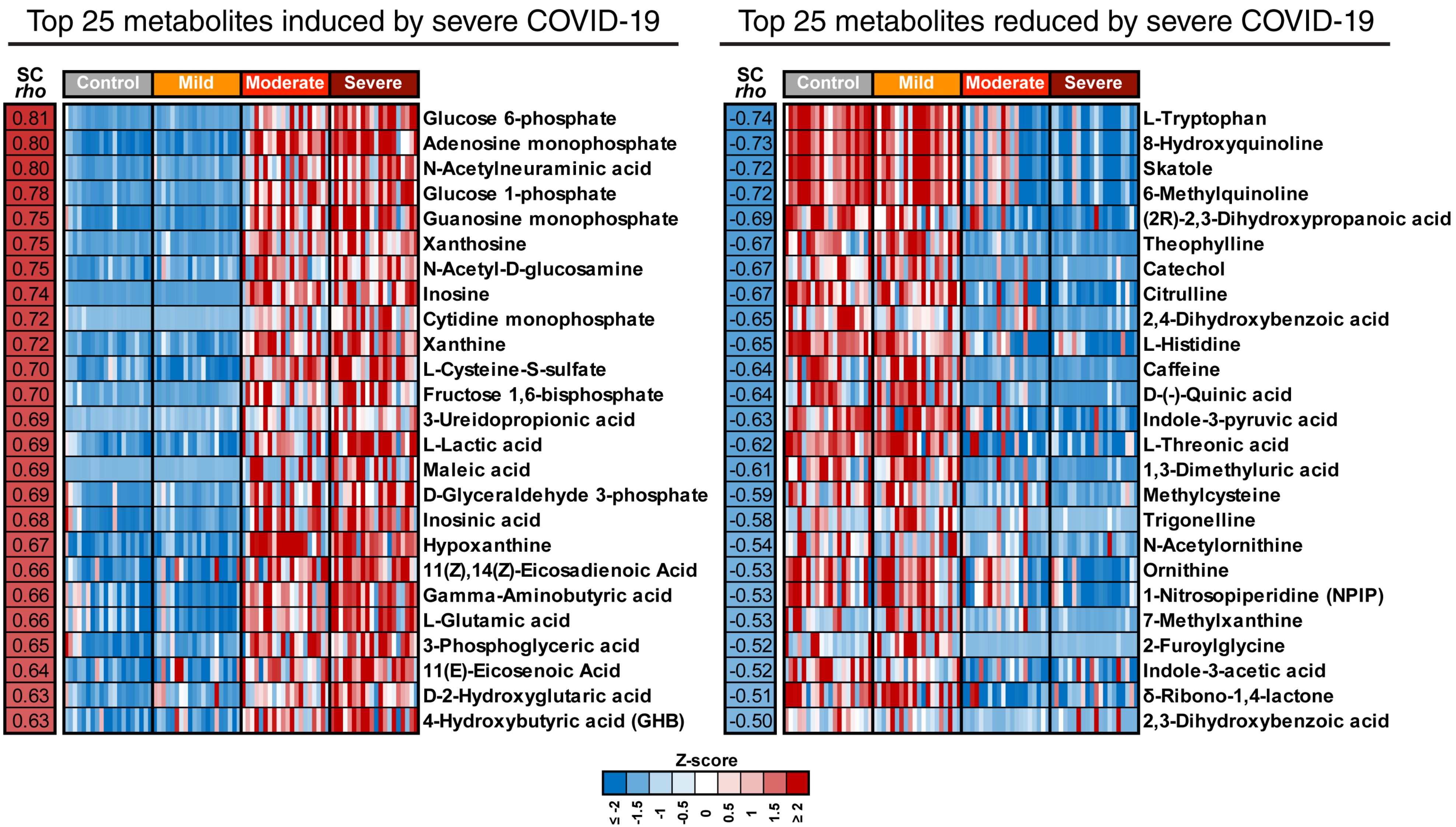

### Supplementary_Figure_3

#### Supplementary Figure 3

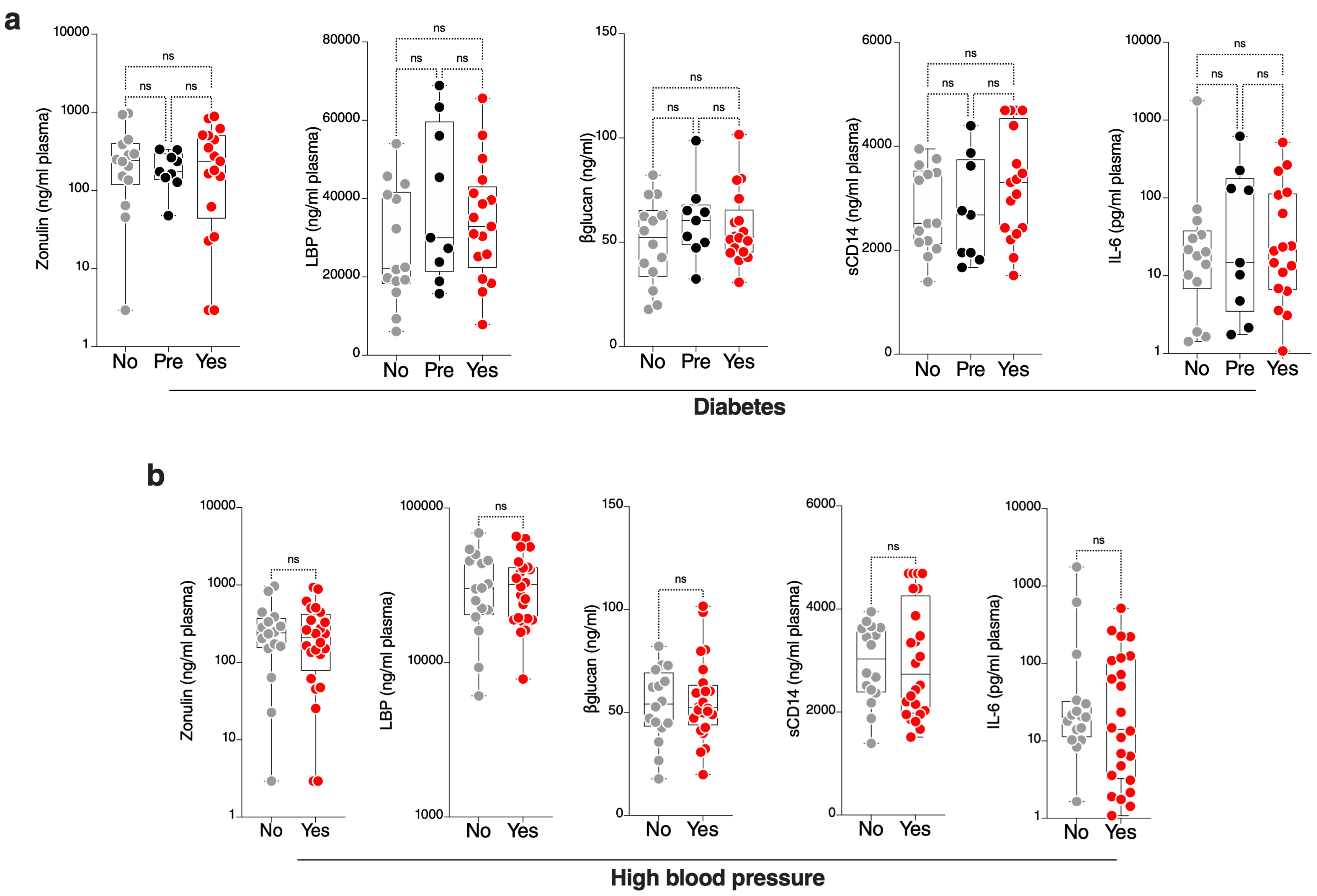
