## Supplementary_Figure_4 for "Plasma Markers of Disrupted Gut Permeability in Severe COVID-19 Patients"

**a**

**Microbial Translocation**

| Glycome | SC *rho* | LBP | Zonulin | sCD14 | βglucan | Occludin | sCD163 |
|---|---|---|---|---|---|---|---|
| SC *rho* | | 0.81 | 0.77 | 0.74 | 0.60 | 0.31 | 0.25 |
| TJAI-binding | 0.74 | 0.66 | 0.57 | 0.56 | 0.42 | 0.21 | 0.11 |
| TcLcl-binding | 0.68 | 0.69 | 0.51 | 0.53 | 0.56 | 0.19 | 0.17 |
| A2 | 0.62 | 0.64 | 0.61 | 0.49 | 0.53 | 0.14 | 0.18 |
| Tetra-sialylated | 0.59 | 0.66 | 0.57 | 0.52 | 0.67 | 0.34 | 0.31 |
| A4G4S4 | 0.59 | 0.66 | 0.57 | 0.52 | 0.67 | 0.34 | 0.31 |
| DSA-binding | 0.52 | 0.45 | 0.33 | 0.40 | 0.51 | 0.10 | 0.11 |
| Tetra-galactosylated | 0.51 | 0.55 | 0.49 | 0.45 | 0.58 | 0.29 | 0.25 |
| ConA-binding | 0.49 | 0.48 | 0.38 | 0.34 | 0.55 | 0.10 | 0.13 |
| Total sialylated | 0.44 | 0.50 | 0.35 | 0.45 | 0.29 | 0.18 | 0.06 |
| AOL-binding | 0.41 | 0.40 | 0.41 | 0.37 | 0.32 | 0.20 | 0.14 |
| Di-sialylated | 0.41 | 0.53 | 0.32 | 0.48 | 0.35 | 0.20 | 0.00 |
| ABA-binding | 0.38 | 0.34 | 0.34 | 0.29 | 0.25 | 0.17 | 0.25 |
| Di-sialylated | 0.36 | 0.32 | 0.45 | 0.24 | 0.29 | -0.02 | 0.02 |
| High-branched | 0.35 | 0.37 | 0.27 | 0.30 | 0.30 | 0.17 | 0.14 |
| AsG2S2 | 0.35 | 0.52 | 0.34 | 0.48 | 0.41 | 0.22 | 0.03 |
| A3F1G3S3 | 0.32 | 0.32 | 0.31 | 0.23 | 0.24 | 0.05 | 0.23 |
| Branched Fucose | 0.31 | 0.30 | 0.30 | 0.22 | 0.22 | 0.04 | 0.22 |
| Fucose | -0.21 | -0.25 | -0.40 | -0.26 | -0.32 | -0.18 | -0.30 |
| AsG2S2 | -0.27 | -0.31 | -0.33 | -0.28 | -0.36 | -0.28 | -0.30 |
| WFA-binding | -0.30 | -0.19 | -0.27 | -0.31 | -0.05 | -0.09 | -0.06 |
| G1FB | -0.33 | -0.31 | -0.26 | -0.12 | -0.32 | -0.12 | -0.12 |
| Core Fucose | -0.33 | -0.47 | -0.32 | -0.48 | -0.31 | -0.20 | -0.08 |
| Terminal di-galactose | -0.35 | -0.29 | -0.26 | -0.33 | -0.22 | -0.26 | -0.22 |
| Low-branched | -0.35 | -0.37 | -0.27 | -0.30 | -0.31 | -0.17 | -0.14 |
| FA2G1 | -0.35 | -0.44 | -0.29 | -0.44 | -0.30 | -0.22 | -0.06 |
| G1F-6_arm | -0.40 | -0.46 | -0.45 | -0.39 | -0.37 | -0.19 | -0.44 |
| G2F | -0.40 | -0.32 | -0.34 | -0.39 | -0.18 | -0.24 | -0.22 |
| A3G3S2 | -0.40 | -0.45 | -0.44 | -0.40 | -0.44 | -0.22 | -0.14 |
| FA2BG1 | -0.42 | -0.51 | -0.38 | -0.34 | -0.38 | -0.09 | -0.19 |
| Non-sialylated | -0.44 | -0.50 | -0.35 | -0.45 | -0.29 | -0.18 | -0.06 |
| Mono-sialylated | -0.45 | -0.49 | -0.39 | -0.47 | -0.41 | -0.32 | -0.10 |
| Termial di-galactose | -0.46 | -0.39 | -0.40 | -0.43 | -0.24 | -0.26 | -0.28 |
| NPA-binding | -0.47 | -0.48 | -0.37 | -0.36 | -0.31 | -0.08 | -0.05 |
| VVA-bindning | -0.47 | -0.42 | -0.40 | -0.49 | -0.19 | -0.28 | -0.15 |
| FA2G2S1 | -0.47 | -0.57 | -0.41 | -0.57 | -0.45 | -0.23 | -0.16 |
| Terminal mono-galactose | -0.49 | -0.59 | -0.39 | -0.53 | -0.51 | -0.31 | -0.06 |
| G2FB | -0.52 | -0.51 | -0.49 | -0.36 | -0.42 | -0.22 | -0.36 |
| FA2G2S1_FABG2S1 | -0.52 | -0.61 | -0.48 | -0.43 | -0.50 | -0.28 | -0.23 |
| M6 | -0.52 | -0.40 | -0.47 | -0.39 | -0.34 | -0.29 | -0.26 |
| FA2G2S1 | -0.53 | -0.67 | -0.45 | -0.48 | -0.60 | -0.09 | -0.23 |
| A2G2_FA2BG1 | -0.53 | -0.58 | -0.42 | -0.38 | -0.40 | -0.22 | -0.11 |
| A4G3S3 | -0.53 | -0.68 | -0.51 | -0.48 | -0.58 | -0.16 | -0.15 |
| FA2BG2 | -0.54 | -0.64 | -0.48 | -0.46 | -0.53 | -0.22 | -0.23 |
| FA2G1 | -0.55 | -0.64 | -0.48 | -0.60 | -0.43 | -0.24 | -0.23 |
| RCA120-binding | -0.56 | -0.54 | -0.49 | -0.48 | -0.44 | -0.33 | -0.10 |
| Terminal galactose | -0.57 | -0.67 | -0.45 | -0.59 | -0.54 | -0.35 | -0.11 |
| UDA-binding | -0.58 | -0.63 | -0.44 | -0.43 | -0.55 | -0.24 | -0.21 |
| ACG-binding | -0.60 | -0.62 | -0.58 | -0.55 | -0.53 | -0.26 | -0.27 |
| STL-binding | -0.60 | -0.50 | -0.43 | -0.39 | -0.46 | -0.08 | -0.06 |
| FA2G2 | -0.66 | -0.73 | -0.54 | -0.70 | -0.50 | -0.30 | -0.25 |
| Terminal di-galactose | -0.69 | -0.77 | -0.57 | -0.68 | -0.56 | -0.30 | -0.25 |

**Cross-correlations**

**b**

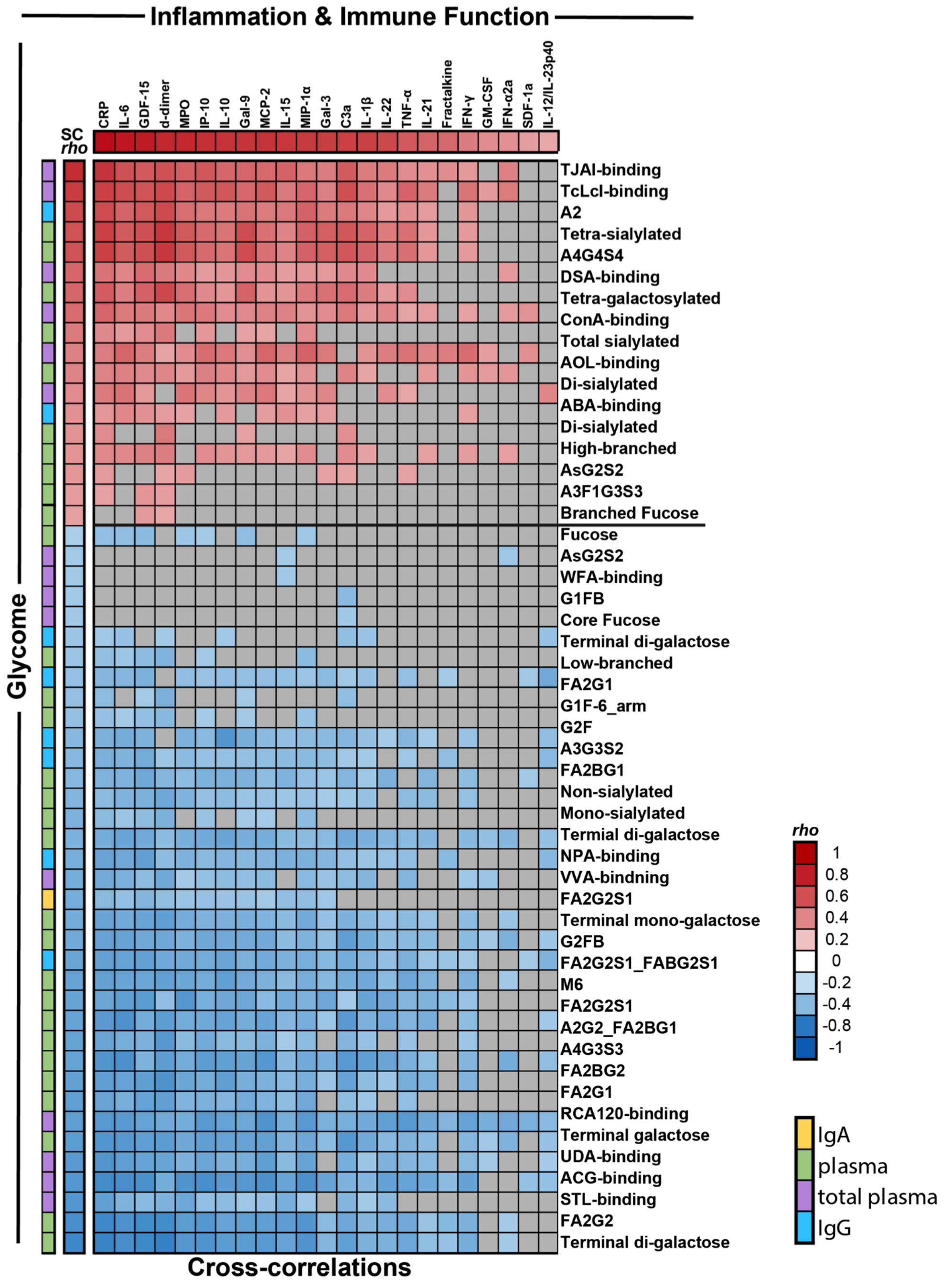
