## Supplementary_Table_1 for "Plasma Markers of Disrupted Gut Permeability in Severe COVID-19 Patients"

**Supplementary Table 1.** Demographic and clinical characteristics of the main study cohort**.**

| | **SARS-CoV-2 Negative -** | **SARS-CoV-2 Positive Mild (outpatients)** | **SARS-CoV-2 Positive Moderate (Inpatients)** | **SARS-CoV-2 Positive Severe (ICU)** |
|---|---|---|---|---|
| Number *(n)* | 20 | 20 | 20 | 20 |
| Female, *n* (%) | 10 (50) | 12 (60) | 11 (55) | 7 (35) |
| Age, years, median (IQR) | 55.5 (15.6) | 52.5 (12.75) | 58.5 (4.25) | 58 (9.25) |
| Deceased, *n* (%) | 0 (0) | 0 (0) | 2 (10) | 6 (30) |
| Body mass index (BMI) | | | | |
| Normal Weight (<25), % | - | 6.7* | 25 | 5 |
| Overweight/obese (>25), % | | 93.3* | 75 | 95 |
| Pre-diabetes, % | - | 6.7* | 15 | 30 |
| Diabetes Mellitus (DM), % | - | 6.7* | 30 | 55 |
| High blood pressure, % | - | 33.3* | 60 | 60 |
| Asthma,% | - | 13.3* | 15 | 5 |
| Hydroxychloroquine, % | - | 6.7* | 40 | 55 |
| Remdesivir, % | - | - | 5 | 25 |
| Tocilizumab, % | - | - | 5 | 30 |
| Chronic Steroid Use, % | - | - | 25 | 5 |
| Acute Steroid Use, % | - | - | 15 | 45 |
| Plasma IV nutrition, % | - | - | 0 | 0 |
| Enteral nutrition use, % | - | - | 100 | 100 |
| Antibiotic administration, % | - | - | 60 | 85 |
| Ethnicity | - | - | | |
| African American, *n* (%) | 0 | 0 | 9 (45) | 7 (35) |
| Hispanic or Latino | 0 | 2 (10.0) | 6 (30) | 9 (45) |
| Caucasian, *n* (%) | 3 (15.0) | 13 (65.0) | 4 (20) | 3 (15) |
| Other, *n* (%) | 2 (10.0) | 1 (5.0) | 0 | 1 (5) |
| Unknown, *n* (%) | 15 (75.0) | 4 (20.0) | 1 (5) | 0 |

*Based on 15 out of 20 donors with available data
