## Supplementary_Table_2 for "Plasma Markers of Disrupted Gut Permeability in Severe COVID-19 Patients"

**Supplementary Table 2.** Demographic characteristics of the validation cohort.

| | | SARS-CoV-2 Negative<br>- | SARS-CoV-2 Positive<br>Mild (outpatients) | SARS-CoV-2 Positive<br>Moderate (Inpatients) | SARS-CoV-2 Positive<br>Severe (ICU) |
|---|---|---|---|---|---|
| Number *(n)* | | 18 | 20 | 18 | 19 |
| Female, *n* (%) | | 12 (67) | 9 (45) | 6 (33) | 5 (26) |
| Age, years, median (IQR) | | 38.5 (19.25) | 60 (10.5) | 64.5 (14.5) | 63 (15.5) |
| Ethnicity | | - | - | | |
| | African American, *n* (%) | 0 (0) | 3 (15) | 6 (33.3) | 2 (10.5) |
| | Hispanic or Latino | 1 (5.6) | 6 (30) | 9 (50) | 11 (57.9) |
| | Caucasian, *n* (%) | 15 (83.3) | 5 (25) | 3 (16.7) | 5 (26.3) |
| | Other, *n* (%) | 2 (11.1) | 3 (15) | 0 | 1 (5.3) |
| | Unknown, *n* (%) | 0 (0) | 3 (15) | 0 | 0 |
