## Supplementary_Table_3 for "Plasma Markers of Disrupted Gut Permeability in Severe COVID-19 Patients"

**Supplementary Table 3.** The structures and names of *N*-glycans identified in plasma by capillary electrophoresis

| Glycan name and structure | Group | Glycan name and structure | Group |
|---|---|---|---|
| 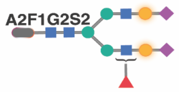 A2F1G2S2 | S2 + ST + FB + LB | 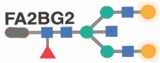 FA2BG2 | G2 + GT + B + FC + LB |
| 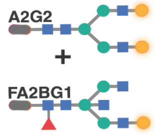 A2G2 + FA2BG1 | G2 + GT + LB | 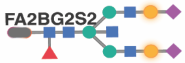 FA2BG2S2 | S2 + ST + B + FC + LB |
| 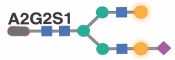 A2G2S1 | S1 + ST + LB | 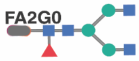 FA2G0 | G0 + FC + LB |
| 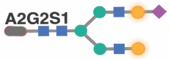 A2G2S1 | S1 +ST + G1 + GT + LB | 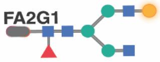 FA2G1 | G1 + GT + FC + LB |
| 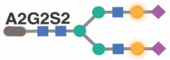 A2G2S2 | S2 + ST + LB | 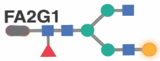 FA2G1 | G1 + GT + FC + LB |
| 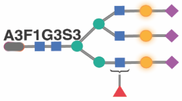 A3F1G3S3 | S3 + ST + FB + HB | 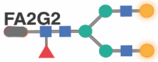 FA2G2 | G2 + GT + FC + LB |
| 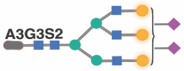 A3G3S2 | S2 + ST + G1 + GT + HB | 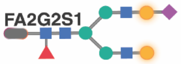 FA2G2S1 | S1 + ST + G1 + GT + FC + LB |
| 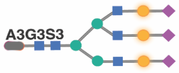 A3G3S3 | S3 + ST + HB | 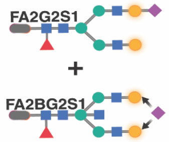 FA2G2S1 + FA2BG2S1 | S1 + ST + G1 + GT + FC + LB |
| 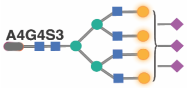 A4G4S3 | S3 + ST + G1 + GT + HB | 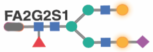 FA2G2S1 | S1 + ST + G1 + GT + FC + LB |
| 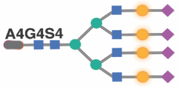 A4G4S4 | S4 + ST + HB | 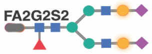 FA2G2S2 | S2 + ST + FC + LB |
| 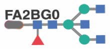 FA2BG0 | G0 + B + FC+ LB | 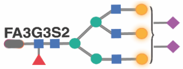 FA3G3S2 | S3 + ST + G1 + GT + FC + HB |
| 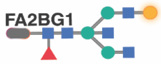 FA2BG1 | G1 + GT + B + FC+ LB | 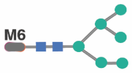 M6 | LB |

▼ Fucose
■ *N*-acetylglucosamine
● Mannose
● Galactose
◆ *N*-acetylneuraminic acid

These 24 glycan structures can be grouped into 15 groups: bisecting GlcNAc (B group), sialic acid (non-sialylated (S0), mono-sialylated (S1), di-sialylated (S2), tri-sialylated and (S3), tetra-sialylated (S4), and total sialylated (ST)), galactose (agalactosylated (G0), mono-galactosylated (G1), di-galactosylated (G2), and total galactosylated (GT)), core fucose (FC group), branched fucose (FB group), high branched (HB group), and low branch (LB group).
