## Supplementary_Table_4 for "Plasma Markers of Disrupted Gut Permeability in Severe COVID-19 Patients"

**Supplementary Table 4.** The structures and names of *N*-glycans identified in isolated IgG by capillary electrophoresis

| Glycan name and structure | Group | Glycan name and structure | Group |
|---|---|---|---|
| A2 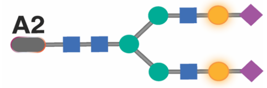 | S2 + ST | G0FB 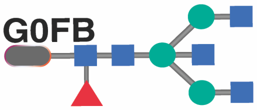 | G0 + F + B |
| A2F 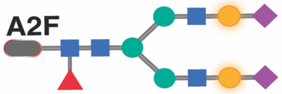 | S2 + ST + F | G1F [6] 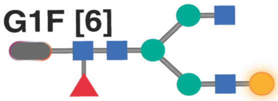 | G1 + GT + F |
| A2FB 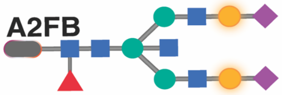 | S2 + ST + F + B | G1F [3] 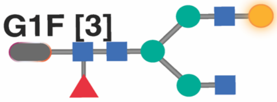 | G1 + GT + F |
| A2B 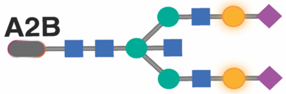 | S2 + ST + B | G1FB 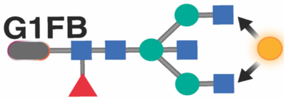 | G1 + GT + F + B |
| G1FS1 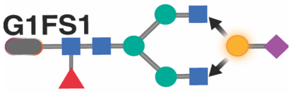 | S1 + ST +F | G0 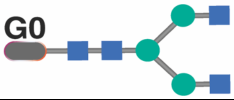 | G0 |
| A1 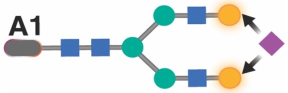 | S1 + ST | G1S1 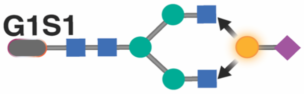 | S1 + ST |
| A1F 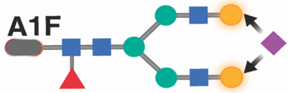 | S1 + ST + F | G2 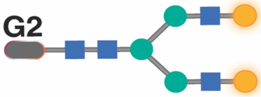 | G2 + GT |
| A1FB 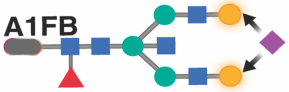 | S1 + ST + F | G2F 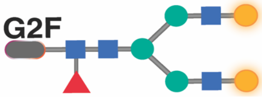 | G2 + GT + F |
| G0F 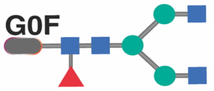 | G0 + F | G2FB 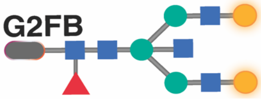 | G2 + GT + F + B |
| G1 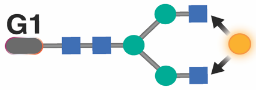 | G1 + GT | G1B [3] 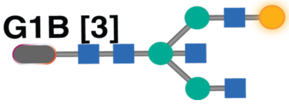 | G1 + GT + B |
| G0B 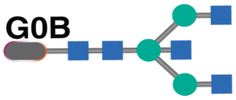 | G0 + B | G1B [6] 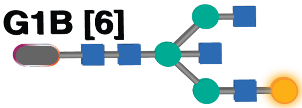 | G1 + GT + B |

- Fucose
- *N*-acetylglucosamine
- Mannose
- Galactose
- *N*-acetylneuraminic acid

These 22 glycan structures were grouped into 9 groups, depending on the presence or absence of four key monosaccharides: bisecting GlcNAc (B group), sialic acid (mon-sialylated (S1), di-sialylated (S2), and total sialylated (ST)), terminal galactose (agalactosylated (G0), mono-galactosylated (G1), di-galactosylated (G2), and total galactose (GT)), and fucose (F group).
