## Supplementary_Table_6 for "Plasma Markers of Disrupted Gut Permeability in Severe COVID-19 Patients"

**Supplementary Table 6.** A list of plasma markers measured in this study.

| Category | Marker | Name | Method of Measurement |
|---|---|---|---|
| **Microbial translocation markers** | Zonulin | haptoglobin 2 precursor | ELISA |
| | LBP | Lipopolysaccharide binding protein | ELISA |
| | β-D-glucan | β-D-glucan | Limulus Amebocyte Lysate (LAL) assay |
| | sCD14 | Soulble CD14 | ELISA |
| | sCD163 | Soulble CD163 | ELISA |
| | OCLN | Occludin | ELISA |
| | I-FABP | Intestinal fatty-acid binding protein | ELISA |
| | Reg3A | Regenerating Family Member 3 Alpha | ELISA |
| **Inflammation and immune function markers** | IL-6 | Interleukin 6 | Multiplex meso scale cytokine assay |
| | TNF-α | tumor necrosis factor alpha | Multiplex meso scale cytokine assay |
| | GM-CSF | Granulocyte-macrophage colony-stimulating factor | Multiplex meso scale cytokine assay |
| | IFN-α2a | interferon α2a | Multiplex meso scale cytokine assay |
| | IFN-β | Interferon beta | Multiplex meso scale cytokine assay |
| | IFN-γ | Interferon gamma | Multiplex meso scale cytokine assay |
| | IL-1β | Interleukin 1β | Multiplex meso scale cytokine assay |
| | IL-2 | Interleukin 2 | Multiplex meso scale cytokine assay |
| | IL-4 | Interleukin 4 | Multiplex meso scale cytokine assay |
| | IL-10 | Interleukin 10 | Multiplex meso scale cytokine assay |
| | IL-12p70 | Interleukin 12 p70 | Multiplex meso scale cytokine assay |
| | IL-12/IL-23p40 | Interleukin 12 p70 | Multiplex meso scale cytokine assay |
| | IL-13 | Interleukin 13 | Multiplex meso scale cytokine assay |
| | IL-15 | Interleukin-12/interleukin 23 p40 | Multiplex meso scale cytokine assay |
| | IL-21 | Interleukin 21 | Multiplex meso scale cytokine assay |
| | IL-22 | Interleukin 22 | Multiplex meso scale cytokine assay |
| | IL-23 | Interleukin 23 | Multiplex meso scale cytokine assay |
| | IL-33 | Interleukin 33 | Multiplex meso scale cytokine assay |
| | Fractalkine | chemokine (C-X3-C motif) ligand 1 (CX3CL1) | Multiplex meso scale cytokine assay |
| | IP-10 | C-X-C motif chemokine ligand 10 (CXCL10) | Multiplex meso scale cytokine assay |
| | MCP-2 | Chemokine (C-C motif) ligand 8 (CCL8) | Multiplex meso scale cytokine assay |
| | MIP-1α | Macrophage inflammatory protein alpha | Multiplex meso scale cytokine assay |
| | SDF-1a | stromal cell-derived factor 1 (SDF1) or C-X-C motif chemokine 12 (CXCL12) | Multiplex meso scale cytokine assay |
| | CRP | C-reactive protein | ELISA |
| | d-dimer | D-dimer | ELISA |
| | MPO | Neutrophil myeloperoxidase | ELISA |
| | GDF-15 | Growth/differentiation factor 15 | ELISA |
| | C3a | Complement component 3a | ELISA |
| | Gal-1 | Galectin-1 | ELISA |
| | Gal-3 | Galectin-3 | ELISA |
| | Gal-9 | Galectin-9 | ELISA |
| **IgG *N*-glycome** | 22 individual glycans structures categorized into 11 groups (Supplementary Table 3) | | Capillary electrophoresis |
| **IgA total glycome** | Binding to 45 lectins with known glycan-binding specificity (Supplementary Table 5) | | Lectin microarray |
| **Plasma *N*-glycome** | 23 individual glycans structures categorized into 20 groups (Supplementary Table 4) | | Capillary electrophoresis |
| **Plasma total glycome** | Binding to 45 lectins with known glycan-binding specificity (Supplementary Table 5) | | Lectin microarray |
| **Plasma Metabolome** | 278 plasma metabolites and three derivative ratios | | Mass spectrometry |
| **Plasma Lipidome** | 2015 plasma lipids categorized into 24 lipid classes | | Mass spectrometry |
