## Supplementary_Table_7 for "Plasma Markers of Disrupted Gut Permeability in Severe COVID-19 Patients"

**Supplementary Table 7.** Top 50 metabolic pathways disrupted by severe COVID-19

| Pathway | *P* value | *FDR* | State | Z | Ratio |
|---|---|---|---|---|---|
| tRNA Charging | 7.9433E-27 | 2.5119E-24 | Activated | 2.828 | 0.22 |
| Superpathway of Citrulline Metabolism | 3.9811E-16 | 7.9433E-14 | | 1.89 | 0.256 |
| Citrulline Biosynthesis | 7.9433E-16 | 1E-13 | | 1.134 | 0.333 |
| Arginine Biosynthesis IV | 3.9811E-14 | 3.1623E-12 | | 0 | 0.333 |
| Purine Nucleotides De Novo Biosynthesis II | 6.3096E-14 | 3.9811E-12 | Activated | 2.121 | 0.22 |
| Superpathway of Methionine Degradation | 1.2589E-11 | 6.9183E-10 | | 0 | 0.127 |
| NAD biosynthesis II (from tryptophan) | 3.1623E-11 | 1.6982E-09 | | 0.816 | 0.226 |
| Asparagine Biosynthesis I | 5.0119E-11 | 2.3442E-09 | | 0.447 | 0.625 |
| Proline Biosynthesis II (from Arginine) | 6.3096E-11 | 2.3988E-09 | | 0 | 0.333 |
| Phenylalanine Degradation IV (Mammalian, via Side Chain) | 6.3096E-11 | 2.3988E-09 | | 0.378 | 0.206 |
| Purine Nucleotides Degradation II (Aerobic) | 7.9433E-11 | 2.7542E-09 | | | 0.2 |
| Urea Cycle | 1.3804E-10 | 3.9811E-09 | | | 0.3 |
| Glycolysis I | 3.3884E-10 | 9.1201E-09 | | | 0.167 |
| L-glutamine Biosynthesis II (tRNA-dependent) | 4.4668E-10 | 1.0965E-08 | | 1 | 0.455 |
| Sirtuin Signaling Pathway | 6.6069E-10 | 1.4454E-08 | | 0.378 | 0.0401 |
| Lysine Degradation V | 6.166E-10 | 1.4454E-08 | | 0.447 | 0.24 |
| Gluconeogenesis I | 7.7625E-10 | 1.5849E-08 | | | 0.149 |
| Alanine Degradation III | 3.7154E-09 | 6.4565E-08 | | 0 | 0.667 |
| Alanine Biosynthesis II | 3.7154E-09 | 6.4565E-08 | | 0 | 0.667 |
| Arginine Degradation VI (Arginase 2 Pathway) | 4.0738E-09 | 6.4565E-08 | | 0 | 0.312 |
| 4-hydroxybenzoate Biosynthesis | 4.0738E-09 | 6.4565E-08 | | 0.447 | 0.312 |
| NAD Biosynthesis from 2-amino-3-carboxymuconate Semialdehyde | 4.0738E-09 | 6.4565E-08 | | 1 | 0.312 |
| Folate Polyglutamylation | 7.9433E-09 | 1.1482E-07 | | -0.447 | 0.278 |
| 5-aminoimidazole Ribonucleotide Biosynthesis I | 7.9433E-09 | 1.1482E-07 | | 1.342 | 0.278 |
| Glutamine Biosynthesis I | 8.7096E-09 | 1.2023E-07 | | -1 | 0.571 |
| Citrulline Degradation | 1.7378E-08 | 2.2909E-07 | | 0 | 0.5 |
| (S)-reticuline Biosynthesis II | 3.0903E-08 | 3.8019E-07 | | 1 | 0.444 |
| Glycine Degradation (Creatine Biosynthesis) | 3.0903E-08 | 3.8019E-07 | | 1 | 0.444 |
| γ-glutamyl Cycle | 3.8905E-08 | 4.6774E-07 | | | 0.208 |
| 4-aminobutyrate Degradation I | 5.1286E-08 | 5.8884E-07 | | 0 | 0.4 |
| Glutathione Biosynthesis | 8.1283E-08 | 8.9125E-07 | | 0 | 0.364 |
| Folate Transformations I | 8.9125E-08 | 9.5499E-07 | | | 0.179 |
| Glutamate Degradation III (via 4-aminobutyrate) | 1.2023E-07 | 1.2589E-06 | | | 0.333 |
| L-carnitine Biosynthesis | 1.7378E-07 | 1.7378E-06 | | -1 | 0.308 |
| Arginine Degradation I (Arginase Pathway) | 1.7378E-07 | 1.7378E-06 | | | 0.308 |
| PFKFB4 Signaling Pathway | 1.9055E-07 | 1.8197E-06 | Activated | 2 | 0.0968 |
| Proline Biosynthesis I | 2.4547E-07 | 2.2909E-06 | | -1 | 0.286 |
| Cysteine Biosynthesis III (mammalia) | 3.8019E-07 | 3.4674E-06 | | | 0.135 |
| Salvage Pathways of Pyrimidine Ribonucleotides | 4.3652E-07 | 3.7154E-06 | | 0 | 0.0609 |
| Selenocysteine Biosynthesis II (Archaea and Eukaryotes) | 4.3652E-07 | 3.7154E-06 | | 1 | 0.25 |
| Citrulline-Nitric Oxide Cycle | 4.3652E-07 | 3.7154E-06 | | | 0.25 |
| Phosphatidylcholine Biosynthesis I | 5.7544E-07 | 4.5709E-06 | | 1 | 0.235 |
| CMP-N-acetylneuraminate Biosynthesis I (Eukaryotes) | 5.7544E-07 | 4.5709E-06 | | | 0.235 |
| Adenine and Adenosine Salvage VI | 6.3096E-07 | 4.8978E-06 | | | 0.6 |
| 4-hydroxyphenylpyruvate Biosynthesis | 6.3096E-07 | 4.8978E-06 | | | 0.6 |
| TCA Cycle II (Eukaryotic) | 7.4131E-07 | 5.4954E-06 | | | 0.119 |
| Uridine-5'-phosphate Biosynthesis | 9.3325E-07 | 6.4565E-06 | | 1 | 0.211 |
| Superpathway of Serine and Glycine Biosynthesis I | 9.3325E-07 | 6.4565E-06 | | | 0.211 |
| Sucrose Degradation V (Mammalian) | 9.3325E-07 | 6.4565E-06 | | | 0.211 |
