## Supplementary_Table_8 for "Plasma Markers of Disrupted Gut Permeability in Severe COVID-19 Patients"

**Supplementary Table 8.** List of the gut-associated and gut microbiota-associated metabolites detected in our study using untargeted LC-MS/MS (50 of the 278 metabolites identified in plasma).

| Name | Reference |
|---|---|
| α-Hydroxyhippuric acid | (Gonthier et al., 2003) |
| β-D-Glucopyranuronic acid | (Pellock and Redinbo, 2017) |
| (2R)-2,3-Dihydroxypropanoic acid | (Monagas et al., 2010) |
| 16-Hydroxyhexadecanoic acid | (Zhitao et al., 2020) |
| 2-Hydroxycinnamic acid | (Coman and Vodnar, 2020) |
| 2-Hydroxyhippuric acid | (Gonthier et al., 2003) |
| 2-Hydroxyvaleric acid | (Skrzypecki et al., 2020) |
| 2,3-Dihydroxybenzoic acid | (Gonthier et al., 2003) |
| 2,4-Dihydroxybenzoic acid | (Marhuenda-Munoz et al., 2019) |
| 3-Hydroxybutyric acid | (Selkrig et al., 2014) |
| 3-Indoxyl sulphate | (Farowski et al., 2019) |
| 3-methylphenylacetic acid | (Marhuenda-Munoz et al., 2019) |
| 4-Hydroxybenzaldehyde | (Marhuenda-Munoz et al., 2019) |
| 4-Hydroxyproline | (Huang et al., 2018) |
| Acetylcholine | (Rea et al., 2016) |
| Allantoin | (Pan et al., 2020) |
| Cholic acid | (Molinero et al., 2019) |
| Choline | (Arias et al., 2020) |
| Citrulline | (Fragkos and Forbes, 2018) |
| D-(-)-Mannitol | (Maekawa et al., 2005) |
| D-Glucose | (Utzschneider et al., 2016) |
| Decanoic acid | (Mathewson et al., 2016) |
| Deoxycholic Acid | (Ridlon et al., 2014) |
| Glucose 6-phosphate | (Di Rienzi and Britton, 2020) |
| Glycine | (Craft et al., 1968) |
| Glycocholic acid | (Ridlon et al., 2014) |
| Glycoursodeoxycholic acid | (Ridlon et al., 2014) |
| Glycyl-L-leucine | (Yasumoto and Sugiyama, 1980) |
| Hippuric acid | (Gonthier et al., 2003;Marhuenda-Munoz et al., 2019) |
| Indole-3-acetic acid | (Gao et al., 2018) |
| Indole-3-lactic acid | (Gao et al., 2018) |
| Indole-3-pyruvic acid | (Gao et al., 2018) |
| Kynurenic acid | (Ridlon et al., 2014) |
| L-Isoleucine | (Krautkramer et al., 2020) |
| L-Kynurenine | (Gao et al., 2018) |
| L-Lactic acid | (Jain et al., 2019;Krautkramer et al., 2020) |
| L-Leucine | (Krautkramer et al., 2020) |
| L-Serine | (Jain et al., 2019;Krautkramer et al., 2020) |
| L-Threonine | (Jain et al., 2019) |
| L-Tryptophan | (Gao et al., 2018;Krautkramer et al., 2020) |
| L-Valine | (Krautkramer et al., 2020) |
| N-Acetyl-DL-tryptophan | (Zhang et al., 2019) |
| Pentadecanoic acid | (Mathewson et al., 2016) |
| Phosphoenolpyruvic acid | (Krautkramer et al., 2020) |
| Pipecolic acid | (Jain et al., 2019) |
| Pyruvic acid | (Krautkramer et al., 2020) |
| Succinic acid | (Oliphant and Allen-Vercoe, 2019) |
| Taurochenodeoxycholic Acid | (Ridlon et al., 2014) |
| Trans-Cinnamic acid | (Kong, 2015) |
| Trimethylamine N-oxide | (Heianza et al., 2020) |
