## Supplementary_Table_9 for "Plasma Markers of Disrupted Gut Permeability in Severe COVID-19 Patients"

**Supplementary Table 9.** Two thousand fifteen lipids identified in this study were assigned to 24 lipid classes.

| Group | Abbreviation | Class | Number of lipids |
|---|---|---|---|
| Phospholipids | CL | Cardiolipin | 2 |
| | LPA | Lysophosphatidic acid | 2 |
| | PA | Phosphatidic acid | 8 |
| | LPC | Lysophosphatidylcholine | 110 |
| | PC | Phosphatidylcholine | 262 |
| | LPE | Lysophosphatidylethanolamine | 22 |
| | PE | Phosphatidylethanolamine | 139 |
| | PG | Phosphatidylglycerol | 10 |
| | LPI | Lysophosphatidylinositol | 6 |
| | PI | Phosphatidylinositol | 47 |
| | LPS | Lysophosphastidylserine | 1 |
| | PS | Phosphatidylserine | 11 |
| Neutral lipids | ChE | Cholesterol ester | 22 |
| | DG/DAG | Diglyceride | 58 |
| | TG/TAG | Triglyceride | 742 |
| Sphingolipids | Cer | Ceramide | 121 |
| | Hex1Cer | Simple Glc series (Ceramide with 1 hexose) | 21 |
| | Hex2Cer | Simple Glc series (Ceramide with 2 hexose) | 10 |
| | Hex3Cer | Simple Glc series (Ceramide with 3 hexose) | 9 |
| | LSM | Lysosphingomyelin | 1 |
| | SM | Sphingomyelin | 359 |
| | SPH | Sphingosine | 1 |
| Other lipids | AcCa | Acyl carnitine | 46 |
| | Co | Coenzyme | 5 |
