## Supplementary material for "Plasma Markers of Disrupted Gut Permeability in Severe COVID-19 Patients": Table_1

**Table 1.** Levels of the 35 out of 50 gut- and gut microbiota-associated plasma metabolites that are disrupted during COVID-19

| | $log_2$ ratio | | | | | | FDR | | | | | |
|---|---|---|---|---|---|---|---|---|---|---|---|---|
| **Group** | Mild | Moderate | Severe | Moderate | Severe | Severe | Mild | Moderate | Severe | Moderate | Severe | Severe |
| **Compared to group** | Control | Control | Control | Mild | Mild | Moderate | Control | Control | Control | Mild | Mild | Moderate |
| 2,3-Dihydroxybenzoic acid | -0.09 | -1.78 | 0.02 | -1.70 | 0.11 | 1.81 | 0.64 | 0.0002 | 0.003 | 0.015 | 0.033 | 0.76 |
| Decanoic acid | -0.04 | -1.68 | -1.32 | -1.64 | -1.28 | 0.36 | 0.90 | 0.004 | 0.009 | 0.0006 | 0.001 | 0.98 |
| (2R)-2,3-Dihydroxypropanoic acid | -0.57 | -1.39 | -2.30 | -0.81 | -1.73 | -0.91 | 0.39 | 0.0005 | $8x10^{-6}$ | 0.004 | $3x10^{-5}$ | 0.78 |
| Indole-3-acetic acid | -0.50 | -1.31 | -0.86 | -0.81 | -0.36 | 0.45 | 0.39 | 0.0002 | 0.001 | 0.007 | 0.023 | 0.92 |
| Pipecolic acid | -0.34 | -1.02 | -1.56 | -0.68 | -1.21 | -0.53 | 0.55 | 0.008 | 0.0002 | 0.24 | 0.012 | 0.48 |
| Hippuric acid | -0.49 | -1.02 | 0.59 | -0.54 | 1.07 | 1.61 | 0.98 | 0.021 | 0.10 | 0.028 | 0.14 | 0.94 |
| 2,4-Dihydroxybenzoic acid | -0.67 | -0.99 | -2.67 | -0.32 | -1.99 | -1.67 | 0.39 | 0.002 | $4x10^{-6}$ | 0.039 | $7x10^{-5}$ | 0.85 |
| Indole-3-pyruvic acid | -0.09 | -0.90 | -1.35 | -0.81 | -1.26 | -0.46 | 0.98 | 0.001 | $1x10^{-5}$ | 0.004 | $9x10^{-5}$ | 0.68 |
| 3-methylphenylacetic acid | -0.52 | -0.75 | -0.77 | -0.23 | -0.25 | -0.02 | 0.64 | 0.031 | 0.013 | 0.56 | 0.84 | 0.92 |
| L-Tryptophan | -0.04 | -0.65 | -0.91 | -0.62 | -0.88 | -0.26 | 0.93 | 0.0001 | $2x10^{-6}$ | 0.0006 | $4x10^{-6}$ | 0.68 |
| Citrulline | 0.04 | -0.57 | -1.08 | -0.61 | -1.12 | -0.51 | 0.95 | 0.002 | $5x10^{-6}$ | 0.003 | $6x10^{-6}$ | 0.51 |
| Deoxycholic Acid | -0.68 | -0.45 | -1.42 | 0.23 | -0.74 | -0.97 | 0.98 | 0.29 | 0.020 | 0.19 | 0.016 | 0.71 |
| Pyruvic acid | 0.03 | -0.37 | -0.52 | -0.40 | -0.54 | -0.15 | 0.96 | 0.020 | 0.001 | 0.013 | 0.0006 | 0.76 |
| L-Serine | -0.36 | -0.30 | -0.61 | 0.06 | -0.25 | -0.31 | 0.39 | 0.079 | 0.001 | 0.78 | 0.18 | 0.51 |
| 4-Hydroxybenzaldehyde | 0.00 | -0.28 | -0.31 | -0.28 | -0.31 | -0.03 | 0.90 | 0.009 | 0.018 | 0.21 | 0.16 | 0.95 |
| L-Threonine | -0.05 | -0.22 | -0.39 | -0.17 | -0.34 | -0.17 | 0.88 | 0.035 | 0.002 | 0.047 | 0.002 | 0.76 |
| 2-Hydroxycinnamic acid | -0.02 | -0.19 | -0.27 | -0.17 | -0.25 | -0.08 | 0.87 | 0.016 | 0.012 | 0.27 | 0.16 | 0.98 |
| Glycine | -0.12 | 0.02 | -0.58 | 0.14 | -0.46 | -0.60 | 0.78 | 0.062 | 0.001 | 0.41 | 0.027 | 0.51 |
| Allantoin | -0.09 | 0.14 | -0.55 | 0.23 | -0.45 | -0.69 | 0.96 | 0.25 | 0.010 | 0.27 | 0.017 | 0.53 |
| 4-Hydroxyproline | -0.04 | 0.29 | 0.11 | 0.33 | 0.15 | -0.18 | 0.90 | 0.020 | 0.67 | 0.018 | 0.93 | 0.48 |
| Glycyl-L-leucine | -0.08 | 0.34 | 0.68 | 0.42 | 0.76 | 0.34 | 0.93 | 0.49 | 0.056 | 0.31 | 0.036 | 0.68 |
| Choline | -0.23 | 0.35 | 0.63 | 0.58 | 0.86 | 0.28 | 0.39 | 0.32 | 0.010 | 0.025 | 0.0003 | 0.68 |
| 2-Hydroxyhippuric acid | -0.98 | 0.64 | 2.42 | 1.61 | 3.40 | 1.79 | 0.49 | 0.075 | 0.028 | 0.73 | 0.24 | 0.76 |
| trans-Cinnamic acid | -0.13 | 0.66 | 0.39 | 0.80 | 0.53 | -0.27 | 0.59 | 0.022 | 0.012 | 0.004 | 0.003 | 0.95 |
| Succinic acid | -0.02 | 0.68 | 1.49 | 0.70 | 1.51 | 0.81 | 0.64 | 0.023 | $7x10^{-5}$ | 0.037 | $2x10^{-5}$ | 0.48 |
| Phosphoenolpyruvic acid | -0.11 | 0.70 | 1.01 | 0.82 | 1.12 | 0.30 | 0.74 | 0.013 | 0.0003 | 0.010 | $7x10^{-5}$ | 0.68 |
| Acetylcholine | -0.02 | 0.72 | 0.55 | 0.74 | 0.57 | -0.17 | 0.69 | 0.028 | 0.053 | 0.012 | 0.036 | 0.92 |
| L-Lactic acid | -0.06 | 0.91 | 1.29 | 0.96 | 1.35 | 0.39 | 0.90 | 0.0002 | $2x10^{-5}$ | 0.0001 | $7x10^{-5}$ | 0.48 |
| L-Kynurenine | 0.28 | 0.96 | 1.20 | 0.69 | 0.93 | 0.24 | 0.39 | 0.002 | $5x10^{-5}$ | 0.024 | 0.002 | 0.68 |
| 16-Hydroxyhexadecanoic acid | 0.94 | 1.46 | 1.19 | 0.52 | 0.25 | -0.27 | 0.39 | 0.0002 | 0.0008 | 0.16 | 0.24 | 0.90 |
| 2-Hydroxyvaleric acid | 1.12 | 1.57 | 1.63 | 0.45 | 0.51 | 0.07 | 0.79 | 0.003 | 0.0004 | 0.025 | 0.016 | 0.94 |
| Glucose 6-phosphate | 0.04 | 1.93 | 2.55 | 1.89 | 2.51 | 0.62 | 0.64 | $2x10^{-5}$ | $3x10^{-6}$ | $1x10^{-5}$ | $4x10^{-6}$ | 0.48 |
| Kynurenic acid | 0.00 | 2.29 | 2.53 | 2.29 | 2.54 | 0.24 | 0.79 | 0.60 | 0.008 | 0.69 | 0.034 | 0.68 |
| ß-D-Glucopyranuronic acid | 0.01 | 2.64 | 2.77 | 2.63 | 2.76 | 0.13 | 0.69 | 0.088 | 0.0002 | 0.27 | $9x10^{-5}$ | 0.68 |
| 3-Hydroxybutyric acid | 0.91 | 2.88 | 3.66 | 1.97 | 2.75 | 0.78 | 0.90 | 0.002 | $5x10^{-6}$ | 0.010 | $7x10^{-5}$ | 0.70 |
