## Supplementary material for "Plasma Markers of Disrupted Gut Permeability in Severe COVID-19 Patients": Table_2

**Table 2.** Logistic regression models of tight junction permeability and microbial translocation markers for their ability to distinguish hospitalized from non-hospitalized individuals.

| Variables | AUC | SE | 95% Confidence interval | | P-value (single predictor models vs. Lasso selected multivariable model) |
|---|---|---|---|---|---|
| Variables identified by Lasso multivariable logistic model | 0.992 | 0 | 0.981 | 1.000 | reference |
| **Zonulin** | 0.951 | 0 | 0.901 | 1.000 | 0.0656 |
| **LBP** | 0.944 | 0 | 0.885 | 1.000 | 0.0987 |
| βglucan | 0.841 | 0 | 0.748 | 0.933 | 0.0008 |
| **sCD14** | 0.930 | 0 | 0.871 | 0.988 | 0.0323 |
| Occludin | 0.663 | 0.1 | 0.542 | 0.783 | <0.0001 |
| Reg3A | 0.635 | 0.1 | 0.510 | 0.759 | <0.0001 |
| sCD163 | 0.627 | 0.1 | 0.503 | 0.751 | <0.0001 |
| IFABP | 0.538 | 0.1 | 0.408 | 0.668 | <0.0001 |

Bold variables are variables selected by Lasso to be included in the multivariable logistic model; AUC = Area under the ROC Curve; SE = Standard error
