## Supplementary material for "Plasma Markers of Disrupted Gut Permeability in Severe COVID-19 Patients": Table_3

**Table 3.** AUC values of the logistic regression model built with data from the main cohort and validated with data from the validation cohort.

| Cohort | n | AUC | SE | 95% Confidence interval |
|---|---|---|---|---|
| **Main** (training) | 79 | 0.9923 | 0.0057 | 0.981 - 1 |
| **Validation** | 75 | 0.886 | 0.042 | 0.803 - 0.968 |
